# Transcriptome deregulation of peripheral monocytes in *GBA*-related Parkinson’s disease

**DOI:** 10.1101/2021.12.22.21267952

**Authors:** Giulietta Maria Riboldi, Ricardo A Vialle, Elisa Navarro, Evan Udine, Katia de Paiva Lopes, Amanda Allan, Madison Parks, Brooklyn Henderson, Kelly Astudillo, Charalambos Argyrou, Maojuan Zhuang, Tamjeed Sikder, Oriol J. Narcis, Shilpa Dilip Kumar, William Janssen, Allison Sowa, Giacomo P Comi, Alessio Di Fonzo, John F. Crary, Steven J Frucht, Towfique Raj

## Abstract

**Background:** Genetic mutations in the beta-glucocerebrosidase (GCase), *GBA* gene, represent the major genetic risk factor for Parkinson’s disease (PD). The function of the *GBA* gene is at the crossroads between the endo-lysosomal pathway and the immune response, two important mechanisms involved in the pathogenesis of PD. However, modifiers of *GBA* penetrance have not yet been fully elucidated.

**Methods:** we characterized the transcriptomic profiles of circulating monocytes and whole blood in a population of patients with PD and healthy controls (CTRL) with (PD/GBA and CTRL/GBA) and without *GBA* variants (iPD and CTRL) (monocytes: n = 56 iPD, 66 CTRL, 23 PD/GBA, 13 CTRL/GBA; whole blood: n = 616 iPD, 362 CTRLs, 127 PD/GBA, 165 CTRL/GBA). Differential expression analysis, pathways enrichment analysis, and outliers detections were performed. Ultrastructural characterization of isolated CD14+ monocytes in the four groups was also performed through electron microscopy.

**Results:** We observed hundreds of differentially expressed genes and dysregulated pathways when comparing manifesting and non-manifesting *GBA* mutation carriers. Specifically, when compared to idiopathic PD, GBA-PD showed dysregulation in genes involved in alpha-synuclein degradation, aging and amyloid processing (i.e. SNCA, LMNA). Gene-based outlier analysis confirmed the involvement of lysosomal, membrane trafficking, and mitochondrial processing in manifesting compared to non-manifesting *GBA*-carriers, as also observed at the ultrastructural levels.

**Conclusions:** Overall, our transcriptomic analysis of primary monocytes identified gene targets and biological processes that can help in understanding the pathogenic mechanisms associated with *GBA* mutations in the context of PD.

## Introduction

Mutations of the *GBA* gene, encoding beta-glucocerebrosidase (GCase), have long been recognized as the major genetic risk factor for Parkinson’s disease (PD) (1–4). Mono- and biallelic mutations of *GBA* can increase the risk of developing PD by 5 to 10 times compared to the general population, with an incidence ranging from 2 to 30% across different ancestries (5). More than 60 pathogenic variants of *GBA* have been identified in PD, where the N370S and L444P mutations account for up to 70-80% (6–8). The molecular mechanisms mainly affected in GBA-PD that have been reported so far encompass the endo-lysosomal pathways, vesicular trafficking, lipid metabolism, and the cell stress response (9–17). Different hypotheses have also been suggested to explain the relationship between *GBA* variants, reduced GCase activity, and alpha-synuclein accumulation, one of the hallmarks of PD (10,14,16). However, because of the incomplete penetrance of the variants of this gene, it is still not clear whether additional modifiers are responsible for the onset of PD in some of the carriers. The identification of these possible modifiers is crucial for targeted therapeutic interventions or to be leveraged as diagnostic biomarkers. In the literature, a possible modifier effect on *GBA* has been reported for variants in the cathepsin B (*CTSB*) and alpha-synuclein (*SNCA*) genes, common variants in the proximity of the *GBA* gene, GBA pseudogene 1 (*GBAP1*), Metaxin 1 (*MTX1*) and Bridging Integrator 1 (*BIN1*) genes, as well as the leucine repeat rich kinase 2 (*LRRK2*), the other major genetic risk factor of PD (18–24).

*GBA* is ubiquitously expressed across tissues and mutations have been associated with aberrant monocyte/macrophage-mediated inflammatory response in both the periphery as well as in terms of microglia activation in the brain of transgenic animal models (25–27). Glial activation appears to start early in the presence of *GBA* variants, as was reported already in asymptomatic carriers of the mutation (28). Since the initial report in 1998 of activated microglial cells in brains of subjects affected with PD, further evidence has stressed a possible role of inflammation in PD pathogenesis involving the peripheral and the central immune response in PD patients (3,29–35). Genetic studies suggest a link between PD and immune response: recent genome-wide association studies (GWAS) identified a number of loci associated with PD which are in close proximity with genes related to the immune and inflammatory response; expression profiles of cells of the innate immune system are also enriched with PD-causative genes; and a polarization of the cis-regulatory effect of common variants associated with PD was identified in the innate immune compartment compared to the adaptive response (36–40). By assessing the transcriptomic profiles of CD14+ monocytes and microglia cells from a large cohort of subjects with PD and healthy controls we were able to describe a distinctive expression profile in these cells, showing dysregulation of genes in the lysosomal and mitochondrial pathways (41).

Because of the described role of the immune system in the pathogenesis of PD and the important role of *GBA* in these cells, monocytes can represent an informative cell-type to assess the role of this mutation in PD (42,43). With the present work we characterized the transcriptomic profiles of isolated CD14+CD16- monocytes in a cohort of PD patients and controls with and without *GBA* mutations and compared its results with expression data from whole blood generated from a validation cohort (Parkinson’s Progression Markers Initiative - PPMI) (**Fig. 1**). We reported a prominent involvement of lysosomal, membrane trafficking, and mitochondrial targets in manifesting vs non-manifesting *GBA*-carriers. Distinctive profiles related to alpha-synuclein-, amyloid- and aging-related processes were detected in manifesting *GBA*-carriers compared to subjects with PD without *GBA* variants. Electron microscopy analysis confirmed the presence of ultrastructural changes in monocytes from manifesting *GBA*-carriers.

**Figure 1.**
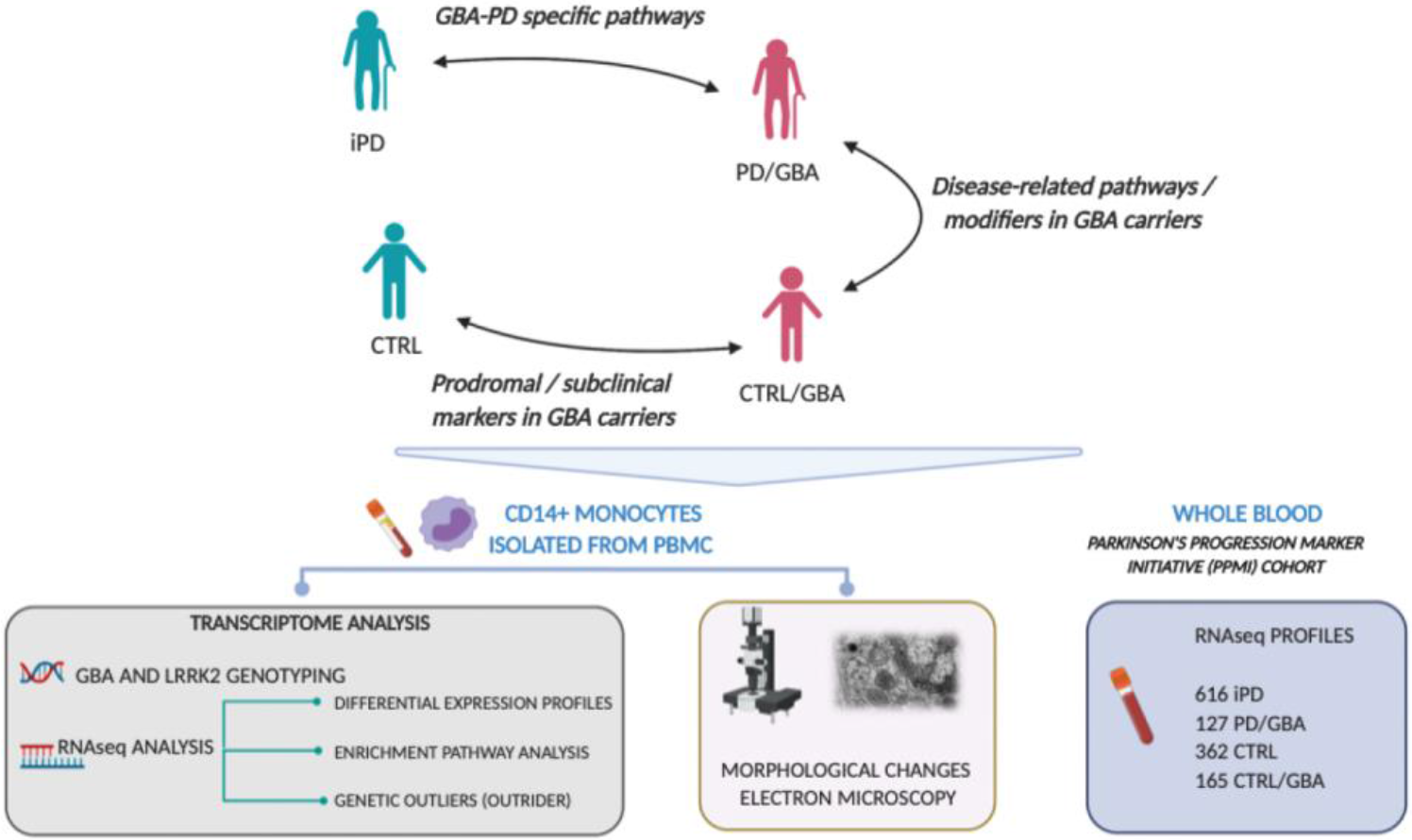
Project design schematic representation. Schematic representation of project design and rationale for the comparison of the selected cohorts and analysis of biological samples. PBMC (Peripheral blood mononuclear cells).

## Results

### Phenotype and genetics drive different transcriptomic profiles in monocytes and whole blood

To characterize the expression profiles of CD14+CD16- monocytes and whole blood in manifesting and non-manifesting *GBA*-carries, we compared the transcriptomic profiles of these cells within the four groups (manifesting *GBA*-carriers (PD/GBA), idiopathic PD patients with no GBA mutations (iPD), non-manifesting *GBA*-carriers (CTRL/GBA), non-manifesting non-*GBA* carriers (CTRL)) from subjects from two independent cohorts: NYMD and PPMI (Fig. 1, Supplementary Table 1). Considering expressed genes with more than 1 CPM in 30% of the samples we found correlation between monocytes and whole blood expressed genes with R = 0.76 (p-value < 0.001) comparing iPD, R = 0.76 (p-value < 0.001) comparing PD/GBA, R = 0.75 (p-value < 0.001) CTRLs, R = 0.75 (p-value < 0.001) comparing CTRL/GBA (Supplementary Fig. 3).

In monocytes, nested analysis based on disease and *GBA*-mutation status showed 512, 1543, and 5 differentially expressed genes (DEG) between PD/GBA and CTRL/GBA, iPD and CTRL, PD/GBA and iPD, respectively, at FDR < 0.05 (Supplementary Fig. 4, Fig. 2a). No DEG at FDR < 0.05 were identified between CTRL/GBA and CTRL subjects (Supplementary Fig. 4). The majority of these genes had a small effect. Considering only genes with a logFC cut-off of 0.5 and FDR < 0.05, we found 84, 57, and 4 differentially expressed genes (DEG) between PD/GBA and CTRL/GBA, iPD and CTRL, PD/GBA and iPD, respectively.

**Figure 2.**
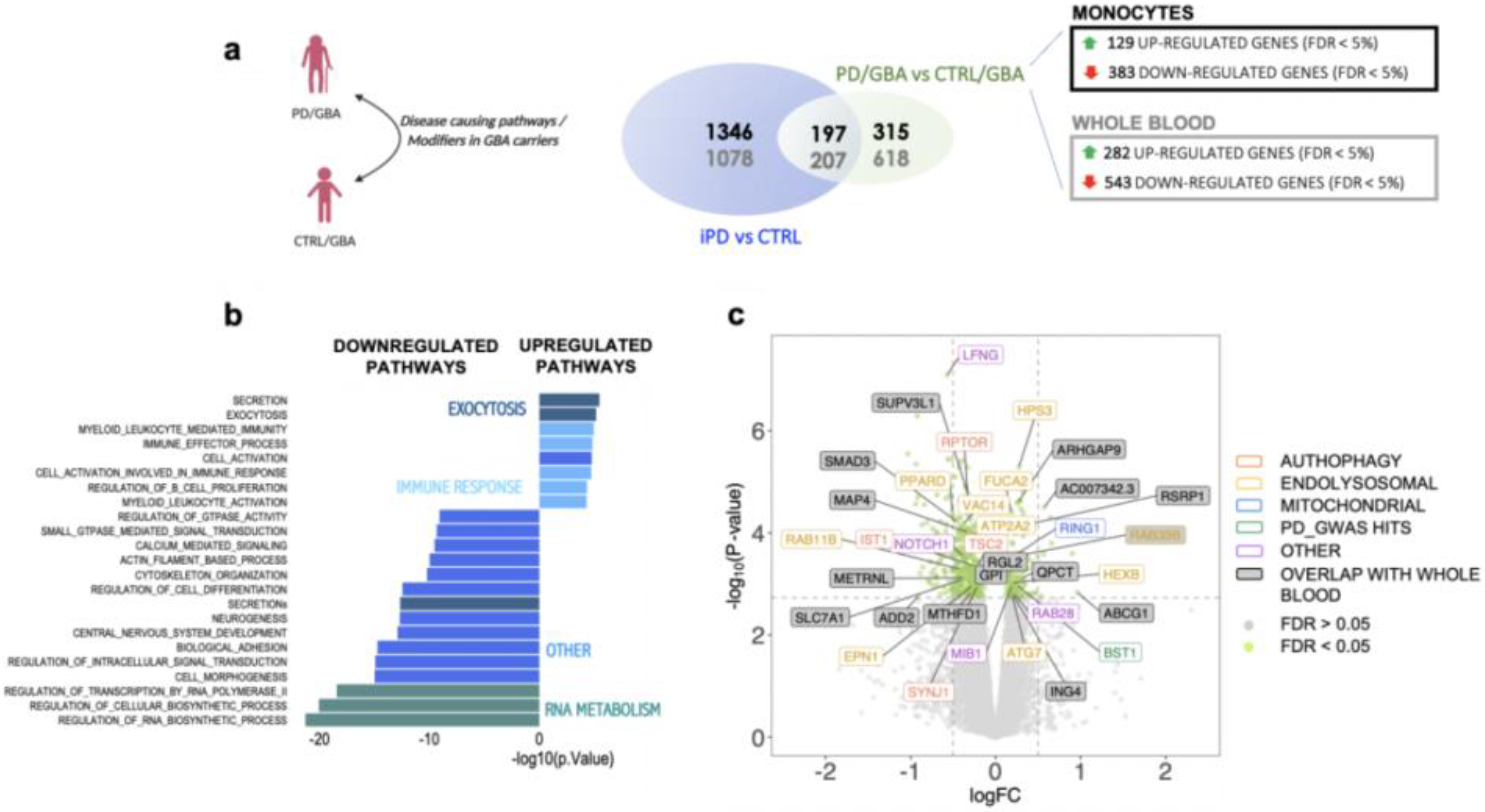
Differential expression profiles between PD and CTRL in GBA carriers. a) Venn-diagram summarizing the number of differentially expressed genes at FDR < 0.05 in monocytes between manifesting and non-manifesting carriers and overlap with genes from non-carriers PD and CTRL subjects (black: data from monocytes, grey: data from whole blood). b) Pathway enrichment analysis of differentially expressed genes between PD/GBA vs CTRL/GBA subjects. Pathway enrichment of genes differentially expressed between PD/GBA and CTRL/GBA subjects with FDR < 0.05 for GO terms are reported. c) Volcano plot represents logFC (x-axis) and −log_10_ of p-values (y-axis) of the differentially expressed genes between PD/GBA and CTRL/GBA groups. Green dots represent genes with FDR < 0.05. Selected genes were highlighted based on overlap with the genes differentially expressed in whole blood (PPMI) and related to targeted pathways as indicated in the legend on the right side.

Transcriptome profiles from whole blood (PPMI cohort) showed 543/282 (upregulated/downregulated), and 549/736 (upregulated/downregulated) DEG in manifesting and non-manifesting *GBA*-carriers, and iPD and CTRLs, respectively at FDR 0.05 (Supplementary Fig. 4).

Interestingly, the level of expression of the *GBA* gene didn’t show any significant differences across the four groups and between PD and CTRL subjects in monocytes or in whole blood (Supplementary Fig. 5a-c). However, lower expression levels of *GBA* were instead observed in carriers of *GBA* variants considered severe, such as the 84GG and V394L (Supplementary Fig. 5a).

### Exocytosis- and myeloid cell activation related genes are impaired in monocytes of manifesting vs non-manifesting GBA carriers

We then further characterized the expression profiles between manifesting and non-manifesting *GBA* mutation carriers as they can potentially elucidate disease mechanisms of *GBA*-related PD. Among the 512 DEG, 197 genes overlapped with genes differentially expressed between iPD and CTRL subjects with no *GBA* mutations in monocytes (**Figure 2A**). 16 genes that dysregulated in monocytes were also differentially expressed in whole blood between manifesting and non-manifesting carriers (Supplementary Table 4). Gene-set enrichment analysis (GSEA) of differentially expressed genes identified a number of pathways. Among the upregulated genes we found enrichment (FDR < 0.05) of pathways related to the myeloid compartment activation, exocytosis, and secretion (Fig. 2b). Downregulated genes were enriched for transcription/RNA-metabolism related pathways, signal transduction (synapses and calcium mediated signal transmission), kinase activity, and vesicle secretion (Fig. 2b). Many of the enriched upregulated and downregulated pathways within monocytes overlapped with enriched pathways of DEG in whole blood (Fig. 2b).

We then aimed to further characterize the involvement of different compartments of membrane trafficking and exocytosis, as well as the lysosomal, ubiquitin, and proteasomal pathways. When we screened for curated pathways and GO pathways related to these terms, we found significant enrichment (adjusted *P-*values < 0.05) only for vesicle membrane within the downregulated genes, which includes genes such as *ATP2A2*, *RIPOR1*, *SYNJ1* (Supplementary Table 2-3). No significant enrichment was identified for the list of membrane trafficking genes from Bandres-Ciga et al., 2019 (9).

Of note, some of the identified DEG are implicated in monogenic forms of PD (such as *ATP13A2* and *LRRK2*), neurodegenerative conditions (such as Tau-related genes), senescence (*FUCA2* and *HEXB)*, and *NOTCH1*-related genes (a highly conserved transmembrane domain protein that is involved in different cellular processes such as cell proliferation, differentiation, apoptotic processes, and modulation of the secretome dynamics in cells) (Fig. 2c, Supplementary Fig. 6a-b). When we assessed the levels of expression of *ATP13A2*, *LRRK2*, and *NOTCH1* in whole blood we were not able to detect significant differences, suggesting the increased power of purified cell population in detecting important changes despite a smaller number of subjects in the cohort (Supplementary Fig. 6c).

Among differentially expressed genes in monocytes, some of which overlap with the pool of differentially expressed in whole blood, we identified targets associated with autophagy (such as *RPTOR*), the endolysosomal, mitochondrial pathways, as well as genes in PD-GWAS loci (Fig. 2c).

We also considered genes that were differentially expressed based on the combined effect of diagnosis and disease status through an interaction model analysis and found DE genes at FDR < 0.15 but no significant hits at FDR < 0.05 (Supplementary Fig. 7). The analysis helped to find genes with a different directionality of expression in the PD/GBA group compared to both the iPD and the CTRL/GBA groups, thus being specifically impaired in the PD/GBA subjects. Among these genes, we identified *FILIP1L* (one of the genes differentially expressed between the PD/GBA and CTRL/GBA groups and that may be coregulated with *LRRK2* and *SNCA* by miRNA-1224) (58), and *HPS3* (Supplementary Fig. 6). *HPS3*, is particularly interesting in this context since this is a gene implicated in the biogenesis of lysosome-related organelles complex-2 (*BLOC-2*). Mutations of this gene are responsible for the Hermansky-Pudlak Syndrome 3. This is a systemic condition affecting the immune system and melanin, similarly to Chediak-Higashi syndrome, previously associated with PD (59).

### Aging- and alpha-synuclein-related pathways are impaired in PD/GBA vs iPD

The number of significantly differentially expressed genes between the subjects with iPD (without *GBA* mutation) and GBA-PD was more limited (Supplementary Fig. 4). We were able to identify 5 genes at FDR < 0.05 (1 upregulated and 4 downregulated) and 44 genes at FDR < 0.15. Pathway enrichment analysis was not possible due to the small number of DEGs between these two groups. However, among the genes with FDR < 0.15 we were able to identify upregulation of the alpha-synuclein gene (*SNCA*) in the PD/GBA group compared iPD subjects, as well as *POLR2D* and *NFATC3*, that are also related to SNCA processing and metabolism (Fig. 3a-b, Supplementary Fig. 8a) (60). We also identified genes related to the amyloid pathways (*ITM2B* and *NCSTN* genes), suggesting an involvement of the aging processes. Interestingly, the most deregulated gene between PD/GBA vs iPD subjects was *LMNA* which is responsible for the progeria syndrome and related to accelerated aging processes (61) (Fig. 3a, b). Few other genes were associated with other known PD causative genes, such as *LMNA* itself (which interact with *LRRK2* on nuclear envelope integrity), *APEX1* (which is degraded by *Parkin* gene) or *MRPL4* (reported to be a rare variant associated with PD) (Fig. 3b) (62–64).Finally, we found a deregulation of *LAMTOR2*, an amino acid sensing and activator of mTORC1 by recruiting it to the lysosome where it is activated (Fig. 3b).

**Figure 3.**
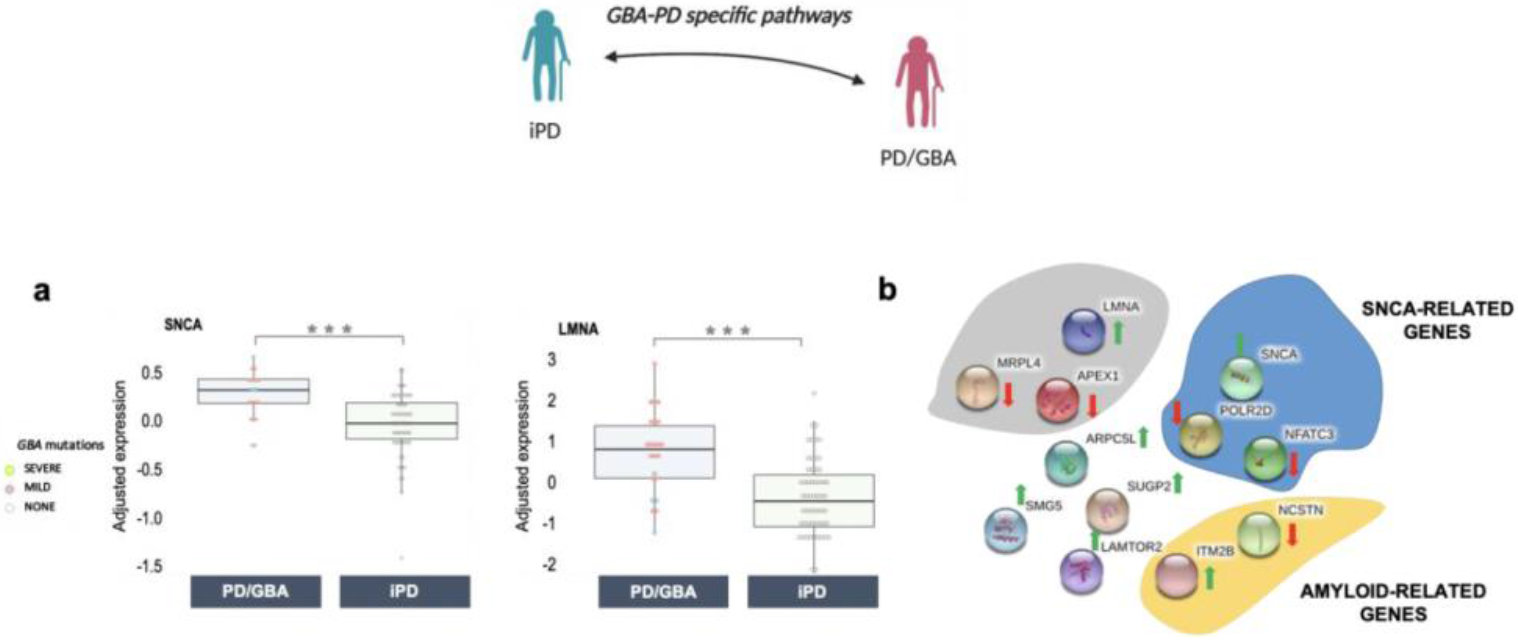
Differential expression profiles of isolated CD14+ monocytes in PD patients with and without GBA variants. a) Differential normalized expression count of *SNCA* and *LMNA* between PD/GBA and iPD. Disease and genetic status are reported on the x-axes. Each dot represents a subject. Dots are colored based on *GBA* mutations (as reported in the legend: *GBA* mild mutations (N370, E326K, R496H), *GBA* severe mutations (L444P/A456P/RecNciI, V394L, 84GG, 84GG/T369M, N370S/RecNciI)). p-value of different expression levels is reported on top (statistics: Mann-Whitney U test). Asterisks indicate significant adjusted p-value (* = adjusted pf-value < 0.05, ** = adjusted p-value < 0.01, *** = adjusted p-value < 0.001). b) Schematic representation (STRING, [150]) of functionally relevant genes differentially expressed between PD/GBA and iPD cohorts. Genes are grouped in colored circles based on shared functional pathways. Arrows indicate whether genes are up (green) or down (red) regulated in the PD/GBA vs the iPD cohort.

Taken together, these observations suggest a deregulation of pathways associated with alpha-synuclein, aging and PD-related genetics in the PD/GBA vs iPD group. When considering differentially expressed genes in whole blood of manifesting *GBA*-mutations carriers and non-*GBA* carriers PD subjects, no significantly differentially expressed genes at FDR < 0.15 were detected, suggesting a more powerful resolution of the analysis in isolated cells, such as monocytes (Supplementary Fig. 4). When considering a nominal p-value < 0.001, some overlapping genes with differentially expressed genes in monocytes were detected, including *LMNA* (Supplementary Fig. 8b-c). Enrichment analysis for differentially expressed genes with nominal p-value < 0.01 highlighted pathways related to the immune response and cellular transport, but not to mitochondria and vesicular trafficking (Supplementary Fig. 8d).

### Gene expression progression patterns of targeted genes

We then decided to follow the progression of the normalized levels of expression in whole blood of some of the targeted genes identified by the above analysis across the three year follow-up between manifesting vs non-manifesting *GBA*-carriers in the PPMI cohort (Supplementary Fig. 9a). Only few of the identified markers showed significant expression across follow-up visits between the two groups, specifically *ING4* and *LRRK2* (Supplementary Fig. 9b). It is important to consider that the number of subjects available in the PPMI cohort for each visit decreased in the two groups across visits (as reported in Supplementary Fig. 9b) which can limit the power of our analysis. It will be worth further validating these results in follow-up data from monocytes in order to assess whether they can be informative biomarker-related pathways in these cohorts.

### Gene expression outliers identified targeted pathways in manifesting and non-manifesting carriers

Gene expression outliers are genes with significantly different levels of expression in a single individual compared to the rest of the population, possibly due to rare genetic variants of those genes in carriers. In order to validate our previous results, we characterized expression outliers between manifesting and non-manifesting carriers. By considering a total of 13,711 genes in 158 total subjects we detected 493 outlier genes and 125 subjects with at least one outlier gene after normalization (Supplementary Fig. 10a-c). No significant enrichment for the number of outlier genes or outlier subjects was detected across our four cohorts after Fisher exact test (% outlier genes per group p-value = 0.479, % outlier subjects per group p-value = 0.456), as well as there was no difference in the proportion of significantly up or down regulated across our cohorts (p-value = 0.06).

Pathway enrichment analysis was performed considering the list of outlier genes in the PD/GBA and the CTRL/GBA group, using GSEA, g-profiler, and IPA. In the PD/GBA group, significant enrichment was detected with g-profiler and IPA for pathways related to Neuroinflammation signaling pathway (including *ICAM1*, *NFATC1*, *IRAK4*), Membrane bounded organelle (such as *VPS41* - involved in vesicular trafficking), ERK/MAPK signaling, and Autophagy (such as *DOCK1* - involved in cytoskeletal rearrangement for phagocytosis of apoptotic cells and mobility of the cells) (Fig. 4a-b).

**Figure 4.**
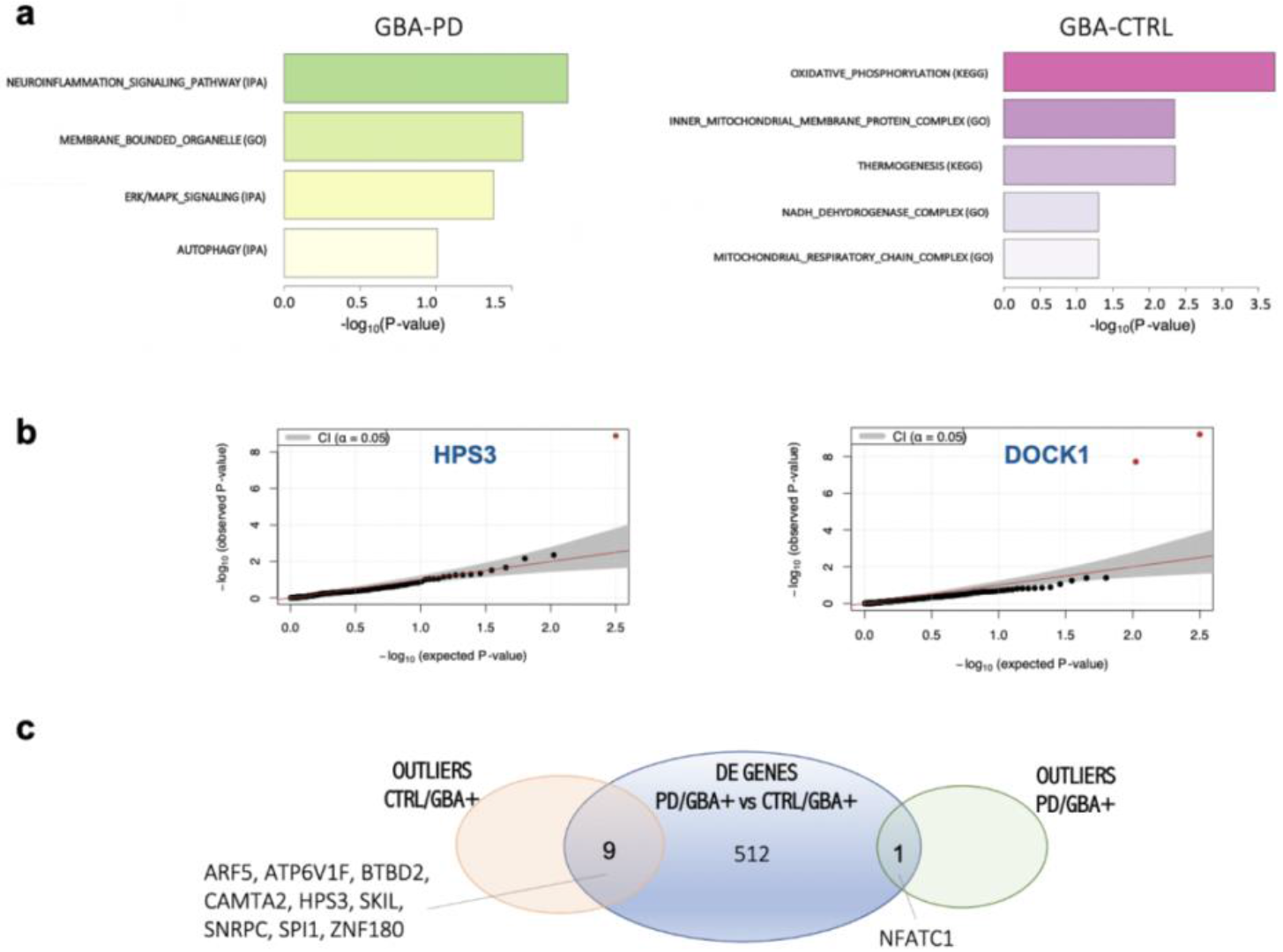
Enrichment analysis of outlier genes in the four cohorts (PD/GBA, CTRL/GBA, iPD, CTRL). a) Pathway enrichment analysis of outlier genes in the PD/GBA and CTRL/GBA cohort (based on GSEA, IPA and g-profiler tools). Bars represent p-value (−log_10_(p-value)). b) Scatter plot representing −log_10_ (p-value) of two of the outliers genes identified in previous analysis. c) Venn-diagram representing overlapping genes between differential expression analysis and outliers analysis in manifesting and non-manifesting carriers in isolated monocytes.

In the CTRL/GBA group pathway enrichment analysis identified the following pathways, all related to mitochondrial functions: Oxidative phosphorylation, Inner mitochondrial membrane protein complex, Thermogenesis, NADH dehydrogenase complex, Mitochondrial respiratory chain complex I. The major driver for pathway enrichment in this group were represented by the following genes: *MT-ATP* (component of the ATP synthase or mitochondrial complex 5, responsible for the final step of oxidative phosphorylation), *MT-ND, MT-ND2, MT-ND4* (all components of the NADH dehydrogenase or mitochondrial complex I) which are all genes encoded by the mitochondrial DNA (Figure 4a, Supplementary Fig. 11). When comparing hits from the outlier analysis and DE between PD/GBA and CTRL/GBA we found 9 genes overlapping the DE data in these two cohorts an in the outliers genes of the CTRL/GBA group and one gene in common with the outliers of the PD/GBA group (*NFATC1*) (Fig. 4b, Supplementary Fig. 11).

### Ultrastructural characterization of CD14+ monocytes confirms membrane vesicle impairment in PD and GBA carriers

Microscopic analysis of CD14+ monocytes from our four cohorts were performed to assess how transcriptomic changes and enriched pathways are reflected at the ultrastructural level. Qualitative assessments showed that in the CTRL/GBA and CTRL groups samples showed normal cell size (12-20 μm), with intact cytoplasm organelles (Golgi (G), endoplasmic reticulum (ER)/rough ER, ribosomes, pinocytotic vesicles, lysosomal granules, phagosomes, mitochondria, and microtubules), indented nuclei, and pseudopodia extend from cell surfaces (Fig. 5a, i/i’; ii/ii’).

**Figure 5.**
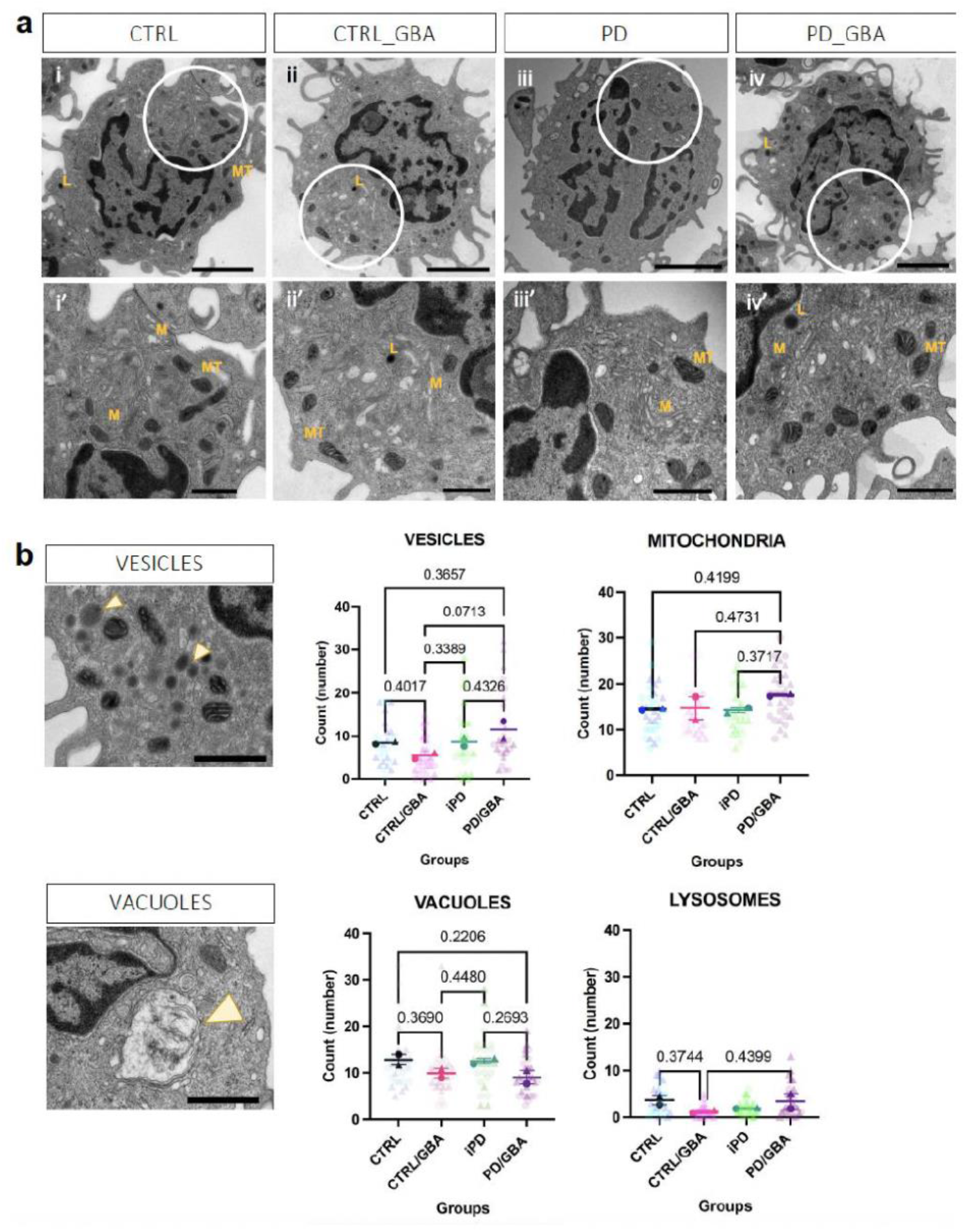
Morphological characterization of CD14+ monocytes. **a)** Low magnification (1800X) images (i, ii, iii, iv) and high magnification (4000x) images (i’, ii’, iii’, iv’). Location of the high magnification images within low magnification pictures are highlighted by the white circle in the first row. Scale bar: 2 **μ**m and 800 nm respectively. i/i’) Cells from CTRL and ii/ii’) CTR/GBA subjects showing normal membrane compartment (M, Golgi and Endoplasmic reticulum), lysosomes (L), mitochondria (MT), cell-to-cell adherences, multiple pseudopodal extensions from the cell membrane. iii/iii’) Cells from iPD subjects showing highly thickened, and distorted cell membrane compartment (M, Golgi and Endoplasmic reticulum); decreased pseudopodia, RER and free ribosomes; mitochondrial (MT) membranes are severely affected, often lacking external membranous encapsulation and with internally swollen cristae. iv/iv’) Cells from PD/GBA subjects showing small and large vacuoles, normal cell membranes, pseudopodal extensions appear normal, and nuclei. Abnormal membrane assembly (M), ultrastructure, and free ribosomes. **b)** Quantification of mitochondria, lysosome, vesicles and vacuoles in four groups (n= 2 subjects per group, 15 cells per sample). Example of vacuole and vesicles reported in the figures on the left highlighted by yellow arrowhead (scale bar 800 nm). Data represents count from each cell from the two samples per group, and mean and SEM of the two replicates. p-value from one-way ANOVA analysis is reported above each comparison (p-value < 0.5 were reported).

Images from the iPD group showed that the majority of cells were relatively intact. However, subpopulations presenting distinct characteristics were identified. In particular, these cells showed impairment of cell membranes, manifesting with enhancing of the cytoplasmic membrane (in the Golgi and ER) and lacking external membranous encapsulation and swelling of internal swollen cristae of the mitochondria (MT) (Fig. 5a, iii/iii’).

In the PD/GBA group, among relatively intact monocytes, distinct subpopulations showed small and large vacuoles, but normal cell membranes, pseudopodal extensions, and nuclei intact. Abnormal membrane-organelles, such as Golgi/ER, RER were severely affected in these cells as well. Quantification for the number of mitochondria, lysosome, vesicles, and vacuolar inclusions didn’t show a statistical difference between the four groups (Fig. 5b).

## Discussion

Our work showed that circulating CD14+ monocytes present distinctive transcriptome profiles, with deregulated molecular pathways and ultrastructural changes linked to PD and variants of *GBA* gene. The involvement of the immune system in the pathogenesis of PD, both of the innate immune system in the periphery and in the brain and the adaptive immune system, has attracted growing attention over the last few years (29–31,33–35,39,65–67). At the same time, GCase, the enzyme encoded by the PD-genetic risk factor *GBA*, is important for the metabolic processes of scavenger cells such as monocytes and macrophages. We found that expression profiles of patients with PD and *GBA* variants were significantly different from the ones of both idiopathic PD patients and non-manifesting carriers (Supplementary Fig. 4) with only a partial overlap with genes differentially expressed between PD and CTRL subjects without *GBA* mutations, suggesting that specific pathways are altered in the presence of variants of this gene (Fig. 2). On the contrary, we didn’t observe significant changes of the expression levels of the *GBA* gene itself (Supplementary Fig. 5). Interestingly, inconsistent results have been previously reported about *GBA* expression levels in the few studies exploring target brain tissues (particularly the substantia nigra) of patients with PD, while reduced levels of GCase activity have been shown in monocytes from patients with PD with and without *GBA* mutations compared to controls (19,68,69).

Deregulated genes and pathways in manifesting *GBA*-carriers compared to the other cohorts highlighted molecular mechanisms previously associated with PD pathogenesis in other cellular or animal models, such as the endo-lysosomal pathway, the mitochondrial pathway, inflammatory markers as well as genes related to monogenic forms of PD and PD GWAS (such as *BST1* and *SNCA*) (Fig. 2) (38,70–72). This suggests that monocytes can mirror a number of processes reported in better characterized cellular and animal models of this disease as well as dopaminergic neurons. Interestingly, transcriptomic signatures were confirmed also at the ultrastructural level. In subpopulations of cells from GBA/PD subjects, we found impairment of the membrane compartment, such as a poor representation of the endoplasmic reticulum and Golgi (Fig. 5). Whether these changes are consistent with an active role of the innate immune compartment in causing the disease or represents just a response to the disease status cannot be inferred at the moment from our data, but it would be worth further exploring. Nevertheless, monocytes may represent a good platform to recapitulate and study PD-associated pathogenic mechanisms. Even more so, despite the small number of subjects in our cohorts – which, to the best of our knowledge, still represents the largest transcriptomic profiles analysis in patients with PD carrying mutations of the *GBA* gene - we were able to identify a large number of differentially expressed genes. This suggests the importance of considering purified cell types, such as isolated CD14+ monocytes, to dramatically reduce the variability due to background noise signals, such as in whole blood.

We also identified deregulation of specific genes related to the endo-lysosomal pathways, such as *ATP13A2* and *LRRK2*, in manifesting *GBA*-carriers (Supplementary Fig. 6). *LRRK2* variants are a common genetic risk factor for the development of PD [56]. In our cohort, expression levels of the *LRRK2* were increased in PD/GBA patients, consistently with the increased activity of this kinase reported in mutated forms associated with PD (73). Previous works supported an interaction between GCase and *LRRK2* (21,22,74). The validation of these results may open the way of considering using the LRRK2-related therapeutic strategies currently tested in ongoing clinical trials also in the large portion of subjects with PD and *GBA* mutations, as well as previously suggested for the *ATP13A2* gene (75). Thus, a growing understanding about the mechanisms concurring to *GBA*-related PD pathogenesis can really contribute to fine-tune therapeutic interventions and can clarify the modulatory effect of these genetic targets on *GBA*.

Impairment of the endo-lysosomal pathways and membrane-trafficking can have a central role in the usual late age of onset in PD, especially in susceptible cells, such as dopaminergic neurons as they undergo a normal process of senescence (72). Interestingly, our analysis identified deregulation of *NOTCH1* and a number of related genes (Supplementary Fig. 6) (76). *NOTCH1* is involved in different molecular pathways in the cells and its turnover and metabolism is very much dependent on vesicle trafficking (77). At the same time, *NOTCH1* regulates aging-related vesicular trafficking, such as aging secretomes, known to accelerate cell degeneration (78). In our analysis, aging-related targets were found to be deregulated when comparing subjects with PD with and without mutations of the gene *GBA* (Fig. 3). The question remains whether aging processes of PD can be further accelerated in carriers of *GBA* mutations or whether the pathogenic variants of this gene are instead responsible in the first place for the activation of pathways that can cause accelerating aging.

Interestingly, we also reported that monocytes of subjects with PD and *GBA* mutations showed an increased expression of the *SNCA* gene, the hallmark protein of PD (Fig. 3). We know that in brains of subjects with *GBA*-related PD there is a robust deposition of this protein, which could explain the more aggressive phenotype in carriers of these mutations compared to idiopathic PD in terms of an earlier age of onset and increased frequency of cognitive impairment and non-motor symptoms (79). It can be speculated that increased expression levels of *SNCA* can represent a compensatory mechanism to its aberrant accumulation due to decreased GCase activity, as previously described (80,81). However, *SNCA* upregulation could also represent a triggered mechanism in predisposed subjects, such as carriers of mutations affecting specific cellular pathways, like *GBA*.

Finally, we identified deregulation of mitochondrial-related genes. In our previous work assessing expression profiles of circulating monocytes and microglia in idiopathic PD patients, we identified an opposite deregulation of the mitochondrial signature in the immune cells in the periphery compared to the central nervous system in PD patients (41). Impaired mitochondrial genes were also detected as outliers among CTRL/GBA subjects, suggesting an involvement and a possible modulatory effect of these genes in the *GBA*-related pathogenesis as previously suggested (Fig. 4) (82).

One limitation of our study, which is shared with many of the works comparing profiles of manifesting vs non-manifesting carriers of mutations with a reduced penetrance, is the fact that we cannot rule out whether some of the non-manifesting carriers of *GBA* mutations will eventually manifest PD at the end of their life. However, because of our large sample size (considering both monocytes and whole blood) and the reduced penetrance of *GBA*, even if some of the subjects who are currently considered non-manifest carriers will eventually phenoconvert to PD, this will not significantly impact the results of our analysis. In the future, it will be interesting to look for the possible prodromal changes in these subjects through longitudinal analysis.

## Conclusions

In conclusion, our results showed that peripheral innate immune cells can be informative in the assessment of disease mechanisms associated with PD. We identified a set of genes and molecular pathways that are specific for *GBA*-related PD, such as a deregulation of SNCA and pathways related to metabolism of beta-amyloid and aging compared to iPD, as well as dysregulation of the lysosomal, membrane trafficking, and mitochondrial processing in manifesting compared to non-manifesting *GBA*-carriers (including genes such as *LRRK2*, *ATP13A2*, *BST1* and *NOTCH1*), also confirmed at the ultrastructural level in monocytes. Further investigation will clarify the possible role as disease biomarkers and of these hits.

## Material and Methods

### Clinical centers and recruitment strategies

Subjects participating in the study were enrolled for the New York Movement Disorder (NYMD) cohort at The Marlene and Paolo Fresco Institute for Parkinson’s and Movement Disorders at the New York University Langone Health (New York), the Bendheim Parkinson Movement Disorders Center at Mount Sinai (BPMD), the Alzheimer’s Research Center (ADRC) and at the Center for Cognitive Health (CCH) at Mount Sinai Hospital (New York). Each Institution’s Institutional Review Board approved the study protocol and the related procedures for subject recruitment, as well as data and samples collection. Subjects were enrolled only upon signing IRB approved informed consent. Enrolled subjects were between the age of 18 and 100 years. The diagnosis of PD was established by qualified movement disorder specialists according to the United Kingdom Parkinson’s Disease Society Brain Bank Clinical Diagnostic Criteria (44). Healthy controls (CTRL) were defined as aged and gender-matched non-affected subjects, who didn’t have a known diagnosis or evidence of PD or other neurological conditions at the time of enrollment. Non-affected subjects were enrolled mostly among participants’ partners and family members.

The study population of PD and CTRL subjects with and without *GBA* mutations was then selected for the study of the transcriptomic profiles of CD14+ monocytes, based on the availability of blood samples, good quality of extracted RNA and sequencing, according to procedures detailed above, as well as self-reported Caucasian ancestry, in order to limit variability due to genetic background architecture related to ancestry. The final population consisted of 56 idiopathic PD (iPD - subjects with PD and without *GBA* mutations), 66 CTRL (non-manifesting subjects without *GBA* pathogenic variants), 23 PD/GBA (subjects with PD and *GBA* pathogenic variants), and 13 CTRL/GBA subjects (non-manifesting subjects with *GBA* pathogenic variants) (Supplementary Table 1). Demographic variables are reported in Supplementary Table 1 and they were accounted for in the downstream normalization of the expression data. The great majority of the selected subjects presented overlap with European ancestry, equally distributed between Ashkenazi Jewish (AJ) and non-AJ ancestry (Supplementary Fig. 1a-b).

### Parkinson Progressive Markers Initiative cohort

Data for the Parkinson Progressive Markers Initiative (PPMI) were downloaded from LONI in January 2021. The PPMI is an international, multicenter, observational study collecting clinical and biological data from subjects with PD, non-affected subjects and cohorts at risk for PD aiming to identify clinically significant biomarkers for the care and diagnosis of PD. Enrolled subjects with PD are *de novo* patients, initially recruited within 2 years from diagnosis. Subjects with PD and controls and known genetic mutations associated with PD, such as *GBA*, *LRRK2*, and *SNCA* are enrolled in the genetic cohort or genetic registry. An extensive description of the PPMI study and collected data and information can be found on www.ppmi-info.org. For this study we selected only subjects with a diagnosis of Parkinson’s disease and non-affected control subjects, with and without mutations of the *GBA* gene. After removing subjects with missing data that would interfere with downstream analysis and considering only baseline visit, the final cohort consisted of 127 PD/GBA, 165 CTRL/GBA, 616 iPD and 362 CTRLs.

### Sample collection and processing

Blood samples were collected fresh on the day of the research visit, in the morning to reduce the variability of sample components and cell activation. Samples were collected in Vacutainer blood collection tubes with acid citrate dextrose (ACD) (BD Biosciences). Samples were processed within 2-3 hours from collection..

DNA was extracted from whole blood (0.5 ml) using the QiAamp DNA Blood Midi kit (Qiagen) according to the manufacturer’s instructions. Nanodrop was utilized to assess DNA quality and concentration.

Sample processing consisted in isolation of peripheral blood mononuclear cells (PBMC) and subsequent CD14+ monocytes purification. For PBMC isolation, SepMate tubes (StemCell Technologies) were used. After dilution in 2-fold PBS (Gibco) tubes were filled with 15 ml of Ficoll-Plaque PLUS (GE Healthcare) and centrifuged at 1,200 g for 10 mins, followed by wash with PBS. Monocyte isolation was performed through sorting of 5 million PBMCs utilizing the AutoMacs sorter with CD14+ magnetic beads (Miltenyi) according to manufacturer’s instructions. Sorted monocytes were stored at −80 °C in RLT buffer (Qiagen) + 1% 2-Mercaptoethanol (Sigma Aldrich). Isolated monocytes stored in RLT buffer were first thawed on ice. RNA was isolated with the RNeasy Mini kit (Qiagen) according to manufacturer’s instructions, including the DNase I optional step. RNA was then stored at −80 °C until library preparation. RNA integrity number (RIN) was assessed with TapeStation using Agilent RNA ScreenTape System (Agilent Technologies). RNA concentration was obtained with Qubit.

For the PPMI cohort, blood samples were centrally collected and processed as reported by the study’s related documentation (www.ppmi-info.org).

### Sample genotyping

Genotyping of the DNA of the samples was performed with the Illumina Infinium Global Screening Array (GSA). This consists of a genome-wide backbone of 642,824 common variants and custom disease SNP content of about 60,000 SNPs. Screening for the most common genetic mutations of the *GBA* and *LRRK2* genes associated with Parkinson’s disease and more frequent among the Ashkenazi Jewish ancestry was performed through targeted genotyping at Dr. William Nichols’ laboratory at the Cincinnati Children’s Hospital. In particular, for the *LRRK2* gene the G2019S variant was screened; for the *GBA* the following 11 variants were analyzed: IVS2+1, 84GG, E326K, T369M, N370S, V394L, D409G, L444P, A456P, R496H, RecNcil. The percentage of each mutation across the entire population and within manifesting and non-manifesting carriers was calculated for both *LRRK2* and *GBA* mutations.

### RNA sequencing

Part of the cohort were processed in house for RNA library preparation. TruSeq Stranded Total RNA Sample Preparation kit (Illumina), with the Low Sample (LS) protocol, was utilized for library preparation according to the manufacturer’s instructions. For the rest of the samples RNA-seq libraries was prepared by a commercial service (Genewiz Inc.). RNA was shipped and processed according to a standard RNA-seq protocol. The Ribo-depletion strategy to remove rRNA was utilized for both samples processed in house and at Genewiz Inc. All samples were sequenced at Genewiz Inc. on an Illumina HiSeq 4000 platform with 150-bp paired-end reads and an average of 60 million reads per sample. Sequencing was performed in four independent batches. For isolated CD14+ monocytes, RNA-seq data were obtained from 56 iPD, 23 PD/GBA, 66 CTRL, and 13 CTRL/GBA subjects.

For the PPMI cohort, RNA was sequenced at Hudson Alpha’s Genomic Services Lab on an Illumina NovaSeq6000. As reported by the PPMI consortium, rRNA+globin reduction and directional cDNA synthesis using the NEB kit were performed. Samples were prepped using the NEB/Kapa (NEBKAP) based library prep, following second-strand synthesis. Sequencing was performed on the Illumina 6000 platform, generating on average 100 million 125 bp paired reads per sample.

### Genotyping and ancestry analysis

Global Screening Array (GSA) was used to genotype each individual. The following quality control metrics were applied: minor allele frequency (MAF) >5%, SNP and samples call rate >95%, Hardy-Weinberg equilibrium (HWE) P-Value > 1 x 10-6. PLINK was utilized to identify and remove duplicated/related samples using pairwise IBD (identity-by-descent) estimation (PLINK PI_HAT values 0.99-1).

Genetic ancestry of samples were confirmed through principal component analysis (45) and comparing multidimensional scaling (MDS) of the values of the study cohort with data from the Phase 3 of 1000 Genome Project samples. For the Ashkenazi Jewish (AJ) only, analyses were repeated using a custom panel as a reference.

### Expression data normalization

FASTQ files were processed with the RAPiD pipeline (46) implemented in the NextFlow framework (RAPiD-nf) (“Nextflow - A DSL for Parallel and Scalable Computational Pipelines” n.d.) providing automated alignment, quantification, and quality control of each RNA-seq sample. To assess quality of the sequences and technical metrics, SAMtools (v1.9) and Picard (2.20) (“Picard Tools - By Broad Institute” n.d.) were utilized prior to and after alignment with FASTQC (0.11.8) (“Babraham Bioinformatics - FastQC A Quality Control Tool for High Throughput Sequence Data” n.d.) (47). Then, reads were processed with trimmomatic (v0.36) for adapter trimming (48). Afterwards, upon creating indexes from GENCODE (v30) (“GENCODE - Human Release 30” n.d.), STAR (2.7.2a) was utilized for aligning the samples to the human reference genome hg38 build (GRCh38.primary_assembly) (49). Quantification of gene expression was obtained with RSEM (1.3.1) (50). Quality control metrics were visualized with MultiQC and gene expression results were generated as raw counts, Counts Per Million (CPM), Transcripts Per Million (TPM), and TMM-voom. The following thresholds were used for initial filtering of the data at the sample level: >20% of reads mapping to coding regions, > 20 Million aligned reads, and ribosomal rate <30%. Sex mismatch was assessed by comparing reported sex with the expression of genes *UTY* and *XIST*, which didn’t identify any sex mismatch in our cohort. At the gene level, genes with <1 count per million in at least 30% of the samples were considered low expression genes and were excluded from the downstream analysis. The above processing led to a total of 13,711 genes that were used in all downstream analyses. Variance partitioning after normalization with surrogate variable (SVs) (explained in paragraph “Linear models for data regression”) showed an average residual less than 25% (Suppl. Fig. 2a). Batch effect seemed to significantly influence data dispersion (Suppl. Figure 2c), that was mitigated after SVs regression (Suppl. Figure 2b, d).

For the PPMI cohort, transcriptome alignment and quantification were performed by the Accelerating Medicines Partnership (AMP)-PD consortium. FASTQ files were aligned using STAR v2.6.1d to the GRCh38 human genome build. Gene quantification was performed using Salmon v0.11.3 using GENCODE v29 annotations. Count per million (CPM) were calculated using edgeR package, and genes with CPM lower than 1 in more than 70% (of a total of 8322 samples from PPMI, PDBP, and BioFIND studies) were removed (total of 18123 remaining genes). Next, only baseline PPMI samples (n=1270) were selected for downstream analysis (only considering patients with PD and control subjects).

### Linear models for data regression

For differential expression analysis, gene counts were scale-normalized by the TMM method using edgeR R package and voom transformed using limma R package (51). Different designs accounting for the majority of available technical and phenotypic variables (rna_batch + Sex + PCT_USABLE_BASES + PCT_RIBOSOMAL_BASES + AJ_gsa_assignment + RIN + PCT_CODING_BASES + PCT_INTERGENIC_BASES + MEDIAN_5PRIME_BIAS + TOTAL_READS + PF_ALIGNED_BASES + PF_MISMATCH_RATE + C1 + C2 + C3 + C4 + C5 + C6 + C7 + C8 + C9 + C10 + PCT_INTRONIC_BASES + PCT_ADAPTER + next_day + C1_AJ + C2_AJ + C3_AJ + C4_AJ + C5_AJ + C6_AJ + C7_AJ + C8_AJ + C9_AJ + C10_AJ + MEDIAN_CV_COVERAGE + PCT_ADAPTER + Diagnosis) were tested. However, principal component and MDS analysis showed the persistence of samples outliers with an impact on the downstream analysis. Therefore, to reduce error rate and increase reproducibility of the data, these were then processed with the ‘sva’ R package for Surrogate Variable Analysis(52). This package allows the identification of surrogate variables to be built directly from a high-dimensional dataset. We estimated 13 surrogate variables while preserving effects from genetic status (presence of *GBA* mutations) and phenotype (subjects with a diagnosis of Parkinson’s disease or control groups). Surrogate variables were built in the design for linear regression. The contribution of known technical and phenotypical variables to the surrogate variables was obtained by linear regression between the surrogate variables and the covariates file and visualized with heatmap.

For the PPMI cohort, the following variables were regressed for downstream analysis: Interaction + age_at_baseline + sex + rin_value + pct_mrna_bases_picard + pct_intergenic_bases_picard + PC1 + PC2 + PC3 + PC4 + PC5 + median_insert_size_picard + plate.

### Differential expression analysis based on interaction between genetic and disease status

A list of differential expressed genes was obtained with the limma package in R by combining expression data (after TMM normalization and voom transformation in R) and surrogate variables. R package limma version 3.38.3 was used to fit a linear model and provide *P*-value upon performing Bayesian moderated t-test. Multiple testing correction with Benjamini-Hochberg False Discovery Rate (FDR) was obtained leveraging the function in the limma package. The cohort of subjects was subdivided into subgroups based on the disease status (subjects with PD vs controls (CTRL)) and *GBA* genetic mutation status (subjects carrying at least one *GBA* variant (PD/GBA, CTRL/GBA) and subjects with no *GBA* variants (iPD, CTRL). We utilized a nested model to analyze expression data in reference to the variable of interest, which consisted in disease status and GBA-mutation status. In addition, to explore the effects of the GBA mutation on the disease status, an interaction term was added to the design as follows: [(PD/GBA – CTRL/GBA) – (iPD – CTRL)]. Results from the comparison of each pair of groups were then extracted. A threshold of FDR < 0.05 was utilized as a threshold of significance.

### Gene set enrichment analysis

Gene set enrichment analysis was performed utilizing the set of differentially expressed genes from the nested interaction model analysis considering genes with FDR < 0.05. Enrichment analysis was performed separately for upregulated and downregulated genes in order to better characterize our set of differentially expressed genes. Gene Set Enrichment Analysis (GSEA) was used for the analysis of different terms from the Gene Ontology (GO) list (specifically: Cellular Component (CC), Molecular Function (MF), and Biological Processes (BP)) (53). Gene-set enrichment with FDR < 0.01 or 0.05 (as specified in the results) were considered. Filters were set for gene-sets with less than 2000 genes. We analyzed up to the first 20 significant enriched terms.

For the dataset obtained from analysis with OUTRIDER (54) tool (see below) the additional following tools were utilized:

1. g-profiler (https://biit.cs.ut.ee/gprofiler/gost), a web server for functional enrichment analysis. Input data were the list of up and downregulated genes separately.
2. Ingenuity pathway analysis (IPA). Canonical data analysis for gene-set enrichment was performed. Statistically significant enriched terms with P-value < 0.05 were accounted for in the final results.

The results from the different tools were then combined together based on P-values after multiple corrections.

### Curated gene-set analysis

Gene-set enrichment analysis was also performed to assess the enrichment in our sets of differentially expressed genes of curated pathways and gene-sets relevant to previously reported pathways related with GBA functions and PD mechanisms. These encompassed genes involved in the following pathways (**Suppl. Table 2**): lysosomal database: 435 genes from The Human Lysosome Gene Dataset; lysosomal storage disease causative gene (LSD list) (54 genes) classified as sphingolipidoses, neuronal ceroid lipofuscinosis, mucolipidosis/oligosaccharides diseases; mitochondrial gene list from (55) (315 genes), classified in distinct mitochondrial pathways as reported in the cited paper, such as mitonuclear cross-talk, mitochondrial dynamics, and OXPHOS; ubiquitin-related gene list (428 genes) from ubiquitin-like modifier activating enzymes and ubiquitin conjugating enzymes E2 (HUGO Gene Nomenclature Committee (HGNC) dataset), and ubiquitin ligase E3. We also assessed targeted enrichment for pathways identified by specific GO and involved in vesicle trafficking and the endolysosomal pathways (based on terms “membrane”, “lysosome”, “endocytosis”, “exocytosis”, that identified 32 pathways, (based on terms “membrane”, “lysosome”, “endocytosis”, “exocytosis”, that identified 32 pathways, Supplementary Table 3). and the following additional pathways: NOTCH1 signaling pathway GO:0007219; senescence associated vacuoles: GO:0010282 (plant); cell signaling via exosome: GO:0099156; cellular senescence: GO:0090398; lipid storage: GO:0019915, GO:0006869; lipid transport GO:0032594; tau protein binding GO:0048156; regulation Tau kinase activity GO:1902947, GO:1902949, GO:1902948; Golgi related pathways: GO:0048211, GO:0005795, GO:0005794, GO:0005796, GO:0051645, GO:0006895, GO:0035621, GO:0055107, GO:0006888. Fisher exact test was run to assess the enrichment of curated terms in the differential expressed gene lists.

### Progression of gene expression levels

Average of gene targeted expression, after normalization, was computed across for four clinical groups (PD/GBA, CTRL/GBA, iPD, CTRL) across 4 times points in whole blood in the PPMI dataset (baseline, year 1, year 2, year 3) and results were plotted to assess the trend of progression of targets selected based on the previous analyses.

### Genetic outliers

RNA-seq data can be also used to identify expression outliers within each single sample that may be expression of underlying genetic mutations, especially in regulatory regions, or compensatory/deregulated mechanisms. Different tools have been reported in the literature to explore this approach, based on Z-score distribution or a combination of Z-scores and the negative binomial distribution, respectively (56,57). These tools presented some limitations such as the lack of specific statistical tests to compare the expression data and the lack of regression for known and unknown covariates that can greatly affect gene expression profiles. OUTRIDER is an additional tool that, instead, utilizes autoencoders to control for variation linked to unknown factors for data normalization. Single genes and single individuals outliers are then detected by comparing univariate cases with the distribution of each gene across the population, by calculating the negative binomial distribution of each single sample compared to all samples (54). Autoencoders are also discharging samples with an excess of outlier genes that may be related to other causes than having a biological relevance (54).

Count per million > 1 in more than 30% of the samples were implemented in the tools. Data were normalized leveraging autoencoders (“OUTRIDER - OUTlier in RNA-Seq fInDER”). Normalized dispersion and mean were then fitted in a binomial model followed by computation of two-sided p-value. The significance threshold was set at an FDR adjusted P-value cut-off of 0.05 and z-score threshold of 2.

### Ultrastructural Characterization of CD14+ Monocytes

CD14+ monocytes were isolated fresh, within 3 hours from collection, from PBMCs as described above. Isolated monocytes were pelleted (*n*=2 per group) from manifesting (PD-GBA), non-manifesting (CTRL-GBA) *GBA* carriers, non-carrier controls (CTRL) and idiopathic PD (iPD). Pellets were fixed (2% glutaraldehyde and 2% paraformaldehyde/ 0.1M Sodium Cacodylate buffer) for one week and Epon resin embedded. Ultrathin sections (75 nm) were collected onto 200 mesh copper grids using a Leica Em UCT ultramicrotome, counter-stained with uranyl acetate/lead citrate, and imaged using a Hitachi 7500 TEM equipped with an AMT digital camera. Images were sized and adjusted for brightness and contrast. Transmission electron microscopy images were acquired for 2-3 cells per field for a total of 15 cells for each sample. Sections were evaluated blinded. Qualitative assessment of the following parameters was assessed and compared among the four different groups: mitochondrial shape, endoplasmic reticulum, lysosomes, nucleus, chromatin, vacuoles, vesicles and inclusions.

### Statistical analysis and graphic representation

For comparison of the different parameters of the ultrastructural characterization between the four groups all analyses were performed with GraphPad Prism version 9.1.0 (GraphPad Inc. La Jolla CA, USA). One-way ANOVA between mean of counts from each sample was performed. Data are presented as Mean and SEM.

For transcriptomic analysis, sample size and statistical methods are reported in the legends of each figure. In the figures, asterisks indicate significant adjusted pvalue (* = adjusted pvalue < 0.05, ** = adjusted pvalue < 0.01, *** = adjusted pvalue < 0.001).

Bioinformatic analysis were performed with R (version 3.6.0) and graphic representation with R Studio version 1.2.1335. Figure 1 was created with BioRender.com.

## Data Availability

All data produced in the present study are available upon reasonable request to the authors

## Declarations

### Ethics approval and Consent to participate

All the procedures involving human subjects were performed upon written informed consent, approval from the institutional review board and in accord with the Helsinki Declaration of 1975. Informed consent was obtained from all individual participants included in the study.

### Availability of data and materials

All data produced in the present study are available upon reasonable request to the authors.

### Authors’ contribution

**Conceptualization:** TR, GMR; **Methodology:** GMR, TR, RAV, EN, EU, KPL, ONJ, SDK, WJ, AS; **Formal analysis and investigation:** GMR, RAV, EN, EU, KPL, AA, MP, BH, KA, CA, MZ, ST, ONJ, SDK, WJ, AS; **Writing - original draft preparation:** GMR, TR; **Writing - review and editing:** GMR, TR, RAV, EN, EU, KPL, AA, MP, BH, KA, CA, MZ, ST, ONJ, SDK, WJ, AA, JFC, ADF, GPC, SJF; **Funding acquisition:** TR, GMR; **Resources:** TR, SJF; **Supervision:** TR.

All authors read and approved the final manuscript.

## Acknowledgements

We thank the study participants at the Marlene and Paolo Fresco Institute for Parkinson’s and Other Movement Disorders for providing their time and participation in the study. We thank Dr. Jack Humphrey for his critical reading of the work. Data used in the preparation of this article were obtained from the Parkinson’s Progression Markers Initiative (PPMI) database (www.ppmi-info.org/data). For up-to-date information on the study, visit www.ppmi-info.org. PPMI – a public-private partnership – is funded by the Michael J. Fox Foundation for Parkinson’s Research and funding partners, including [list the full names of all of the PPMI funding partners found at www.ppmi-info.org/fundingpartners]. G.R. was supported by the American Parkinson’s Disease Association Post-Doctoral Fellowship 2018, the Parkinson’s Foundation Clinical Research Award PF-CRA-1940, and The Fresco Institute for Parkinson’s and Movement Disorders Clinical Fellowship. T.R. supported by grants from the Michael J. Fox Foundation (Grant #14899 and #16743), US National Institutes of Health NIH NINDS R01-NS116006, NINDS U01-NS120256, NIA R01-AG054005, NIA R21-AG063130, and NIA U01 P50-AG005138.

---

**Supplementary Fig. 1.**
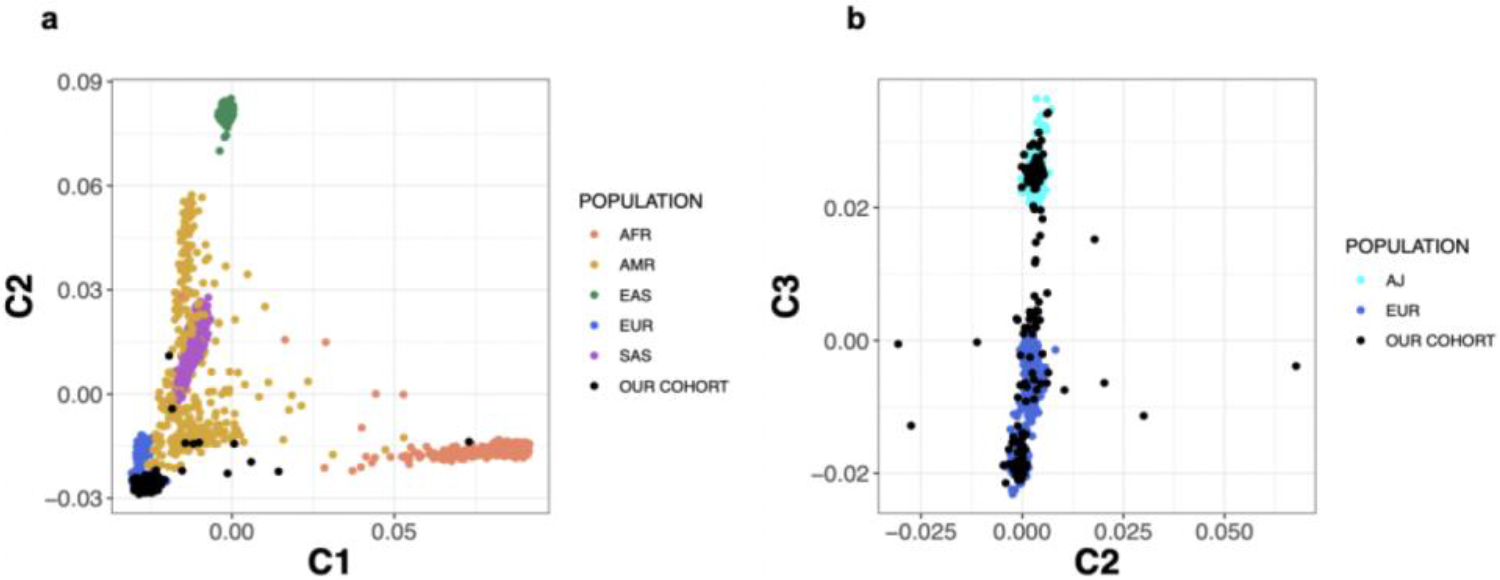
Characterization of genetic background of donor population. a) Representation of PCA analysis of ancestry of MDS values from the cohort of 158 subjects (PD/GBA, iPD, CTRL/GBA, CTRL) compared to 1000 Genome Project samples (Phase 3). The different ancestry are represented in distinct colors (Orange: African; Gold: Ad Mixed American; Green: East Asian; Blue: European; Purple: South Asian; Black: study cohort). B) PCA considering only overlap of MDS values of donor cohort (black) with European ancestry (blue) and AJ ancestry (light blue).

**Supplementary Figure 2.**
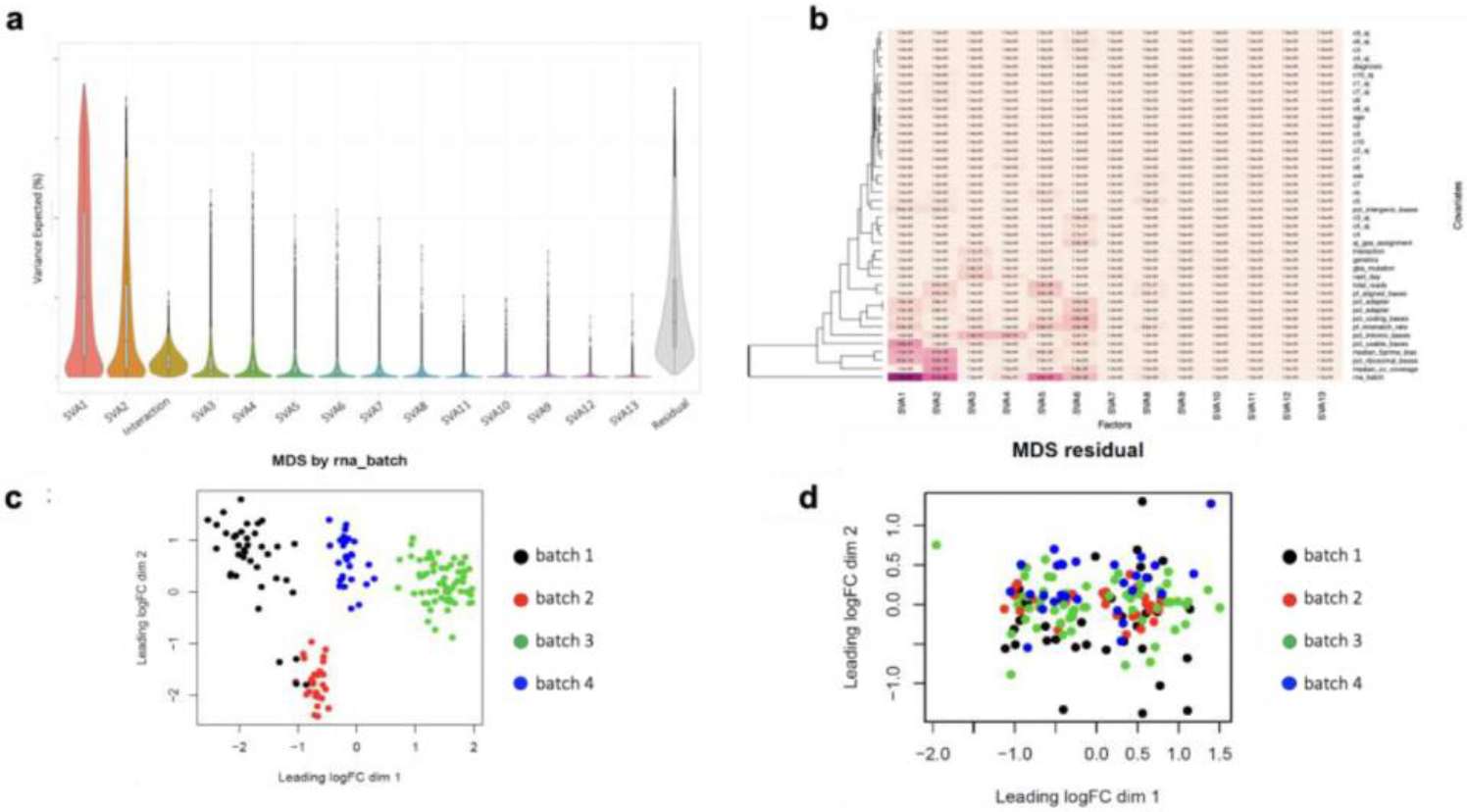
Normalization and quality control of RNA-seq data from isolated CD14+ monocytes. a) Violin plot representing the contribution of each of the surrogate variables (as explained in the text) to the variability of expression data of the study cohort and residual (158 subjects). b) Heatmap representing the results of linear regression between the surrogate variables utilized for data normalization and technical variables (from RNA-seq analysis) and metadata. Coefficient of linear regression is reported in the heatmap for each correlation pair. c) Distribution of MDS values of study cohort identified a clear clustering based on batches used for RNA-seq analysis (batches 1 to 4). d) After regression of SVs, variability of MDS values is significantly reduced, with no significant outliers and no clustering based on experimental batches.

**Supplementary Fig. 3.**
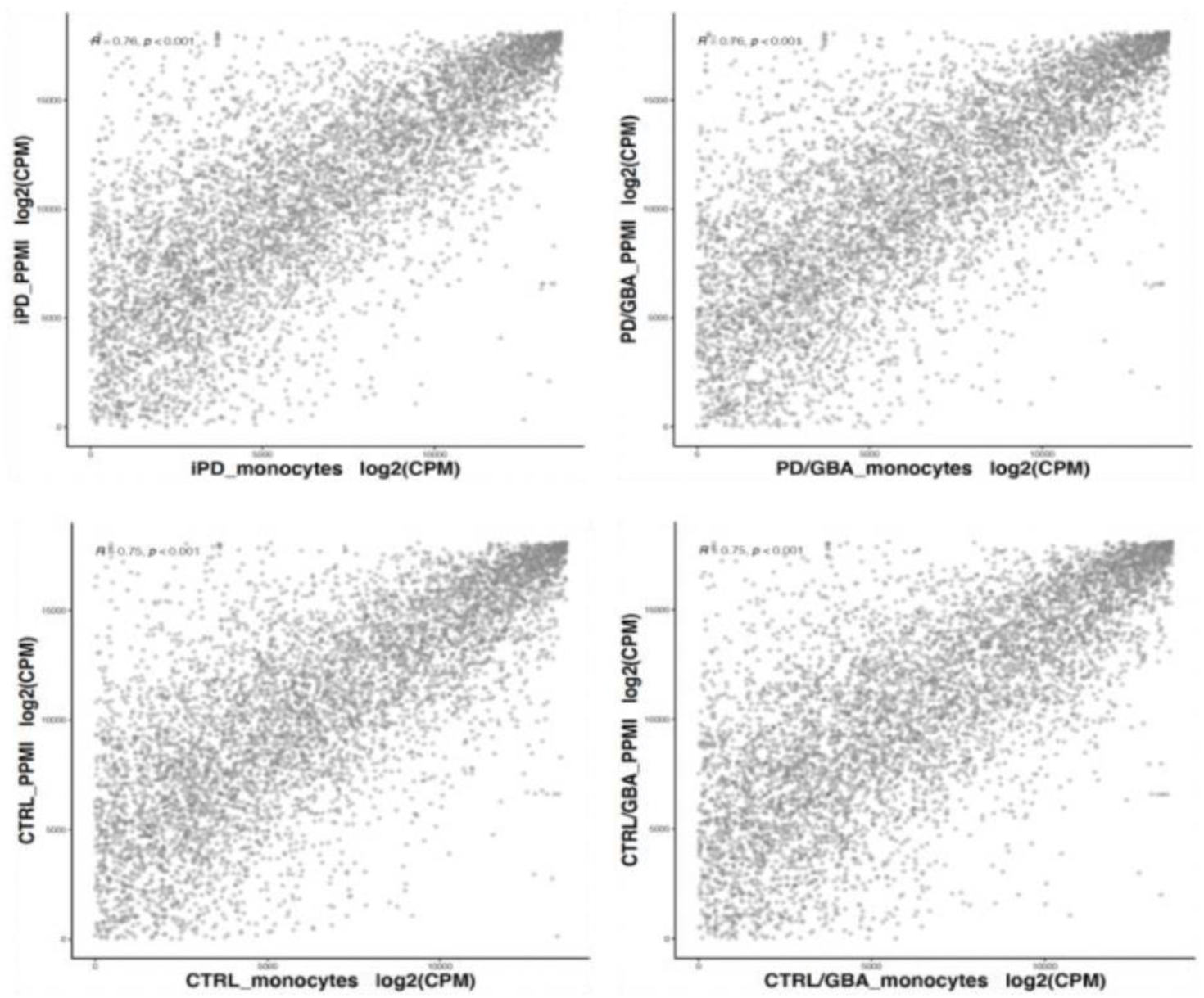
Correlation between gene expression levels in isolated CD14+ monocytes and whole blood. Genes with expression with more than 1 CPM in 30% of the samples were considered from both cohorts (discovery cohort: isolated CD14+ monocytes, validation cohort: whole blood - PPMI cohort). Spearman correlation between levels of normalized mean gene expression across subjects within each cohort per sub-group of subjects was calculated (iPD - top left R = 0.76 p < 0.001, PD/GBA - top right R = 0.76 p < 0.001, CTRL - bottom left R = 0.75 p < 0.001, CTRL/GBA - bottom right R = 0.75 p < 0.001).

**Supplementary Fig. 4.**
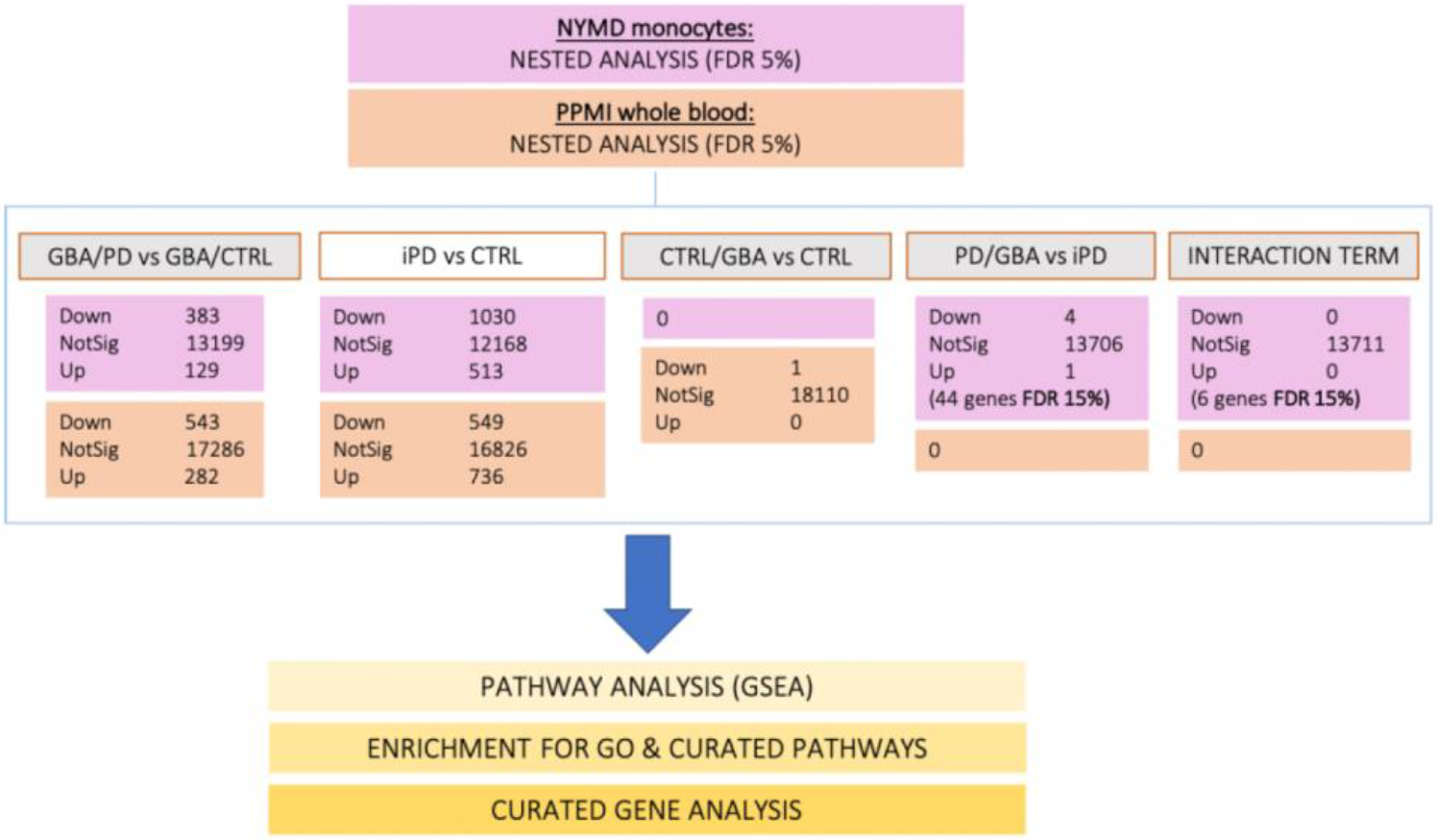
Differential expression analysis of transcriptomic data from purified CD14+ monocytes and whole blood. Summary of the number of differentially expressed genes across the four groups (iPD, PD/GBA, CTRL, CTRL/GBA) in monocytes (pink boxes) and whole blood (orange boxes). The number of upregulated (Up), downregulated (Down) and not-significantly deregulated genes at FDR < 0.05 (or otherwise specified in the box) are reported for each comparison in the below boxes. Differentially expressed genes were analyzed to study pathway enrichment analysis with the listed tools (GSEA, gprofiles, IPA), as well as for enrichment of curated pathway analysis and curated gene analysis (as reported in yellow boxes).

**Supplementary Fig. 5.**
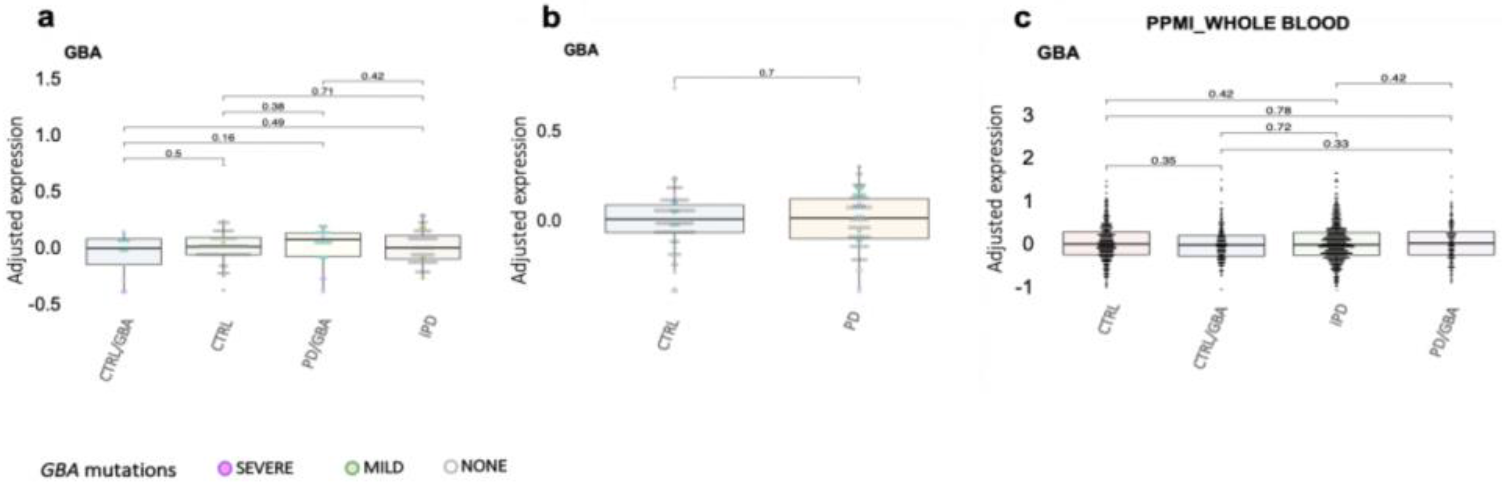
Differential expression of *GBA* in CD14+ isolated monocytes and whole blood. Box plot representing differential expression levels (normalized expression count) of *GBA* in isolated CD14+ monocytes (a and b) and whole blood (c). In b) data from isolated CD14+ monocytes of *GBA*-carries and non-carriers within PD and CTRL subjects were combined and compared. Each dot represents a subject. Dots are colored based on *GBA* mutations (as reported in the legend: *GBA* mild mutations (N370, E326K, R496H), *GBA* severe mutations (L444P/A456P/RecNciI, V394L, 84GG, 84GG/T369M, N370S/RecNciI)). p-value of different expression levels is reported on top (statistics: Mann-Whitney U test). Asterisks indicate significant p-value (* = p-value < 0.05, ** = p-value < 0.01, *** = p-value < 0.001).

**Supplementary Fig. 6.**
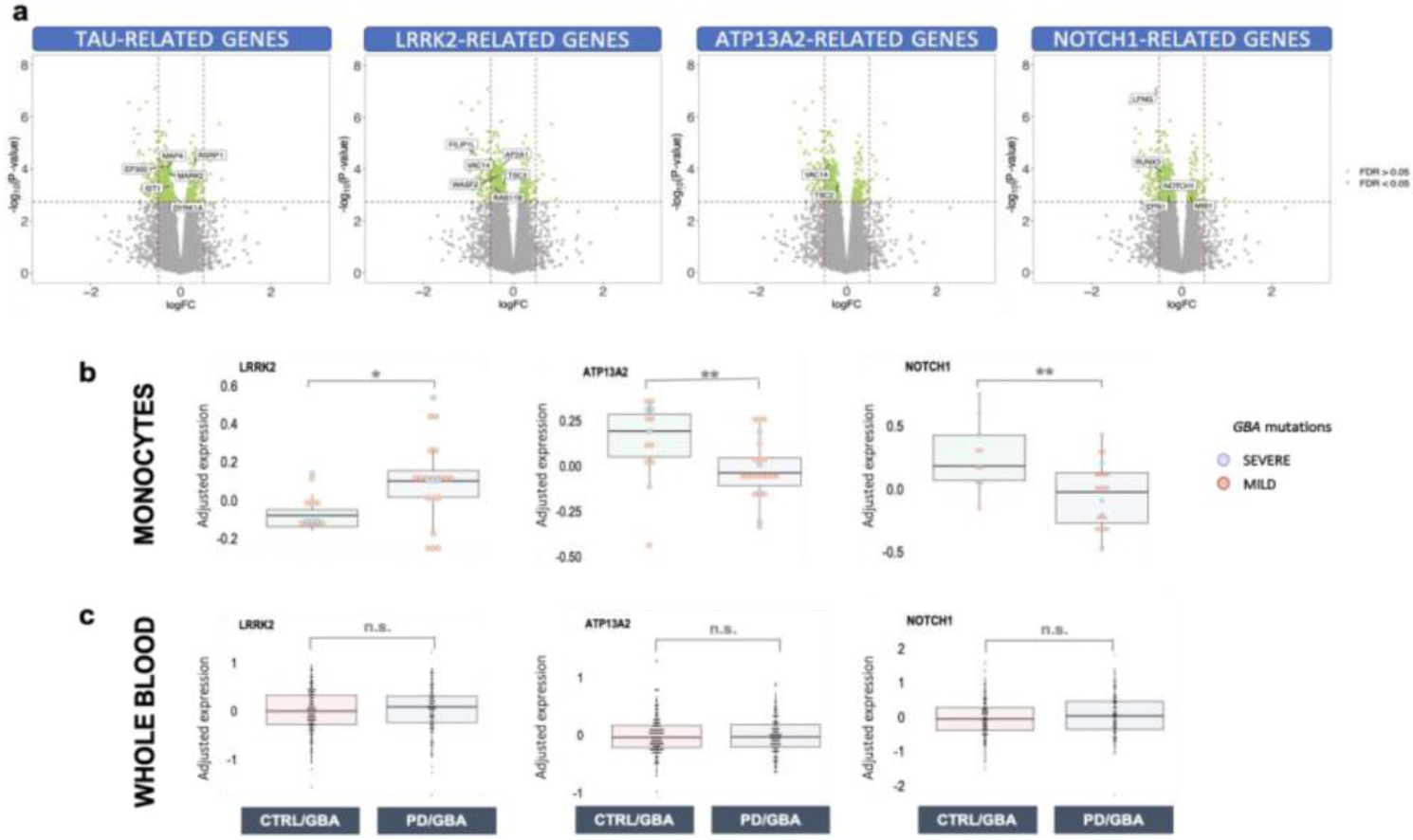
Differential expression of target genes in manifesting and non-manifesting carriers. a) Volcano plot showing differentially expressed genes with FDR < 0.05 (green dots) in isolated monocytes from manifesting vs non-manifesting carriers. Genes related to targeted pathways (*LRRK2*, *ATP13A2*, *NOTCH1* and *TAU*) are highlighted. Differential levels of expression of the targeted genes (*ATP13A2*, *LRRK2*, *NOTCH1*, between manifesting and non-manifesting carriers in isolated monocytes (b) and whole blood (c) from manifesting and non-manifesting GBA-mutation carriers. Each dot represents a subject. Dots are colored based on *GBA* mutations (as reported in the legend: *GBA* mild mutations (N370, E326K, R496H), *GBA* severe mutations (L444P/A456P/RecNciI, V394L, 84GG, 84GG/T369M, N370S/RecNciI)). p-value of different expression levels is reported on top (statistics: Mann-Whitney U test). Asterisks indicate significant p-value (* = p-value < 0.05, ** = p-value < 0.01, *** = p-value < 0.001).

**Supplementary Fig. 7.**
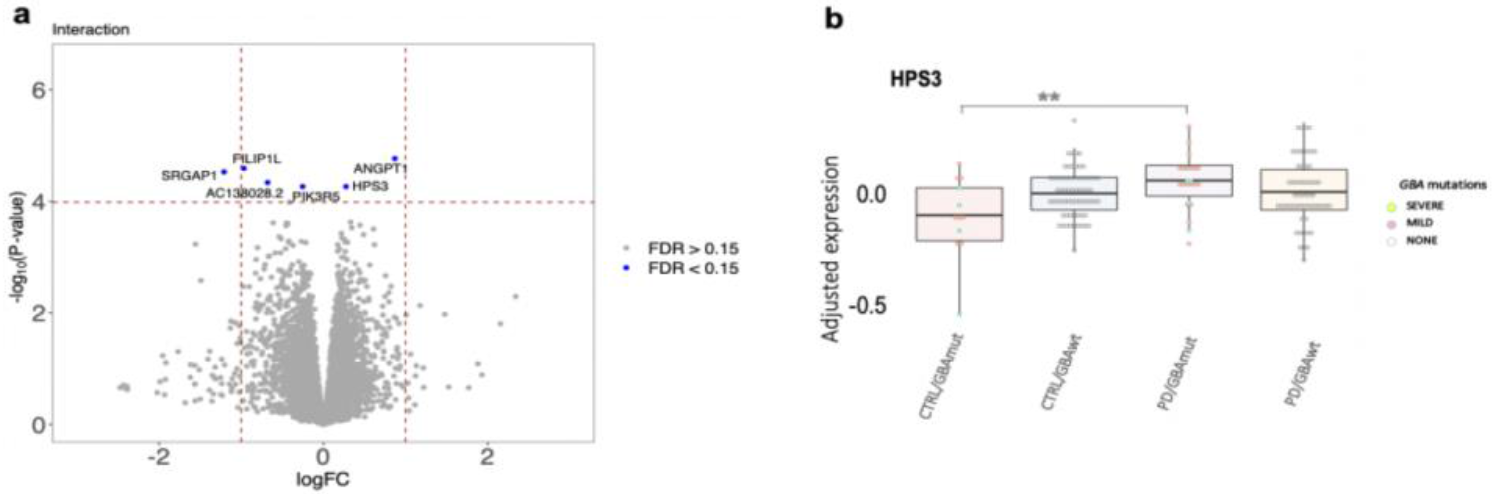
Differential expression profiles across the four cohorts based on diagnosis and genetic status interaction. a) Volcano-plot representing log_2_ fold change (x-axes) and P-Value (y-axes, -log_10_ P-Value) of differential expressed genes based on diagnosis and genetics interaction between the four cohorts (PD/GBA, CTRL/GBA, iPD, CTRL/GBA). Genes with FDR < 0.05 are highlighted in blue and labeled with their IDs. Differentially expressed genes encompassed: *ANGPT1* (angiopoietin gene involved in angiogenesis), *FILIP1L* (Filamin A Interacting Protein 1 Like), *AC138028.2* (novel transcript), *SRGAP1* (Slit-Robo GTPase-activating protein 1), *PIK3R5* (Phosphoinositide 3-kinase regulatory subunit 5, responsible for Ataxia with Oculomotor-Apraxia type 3), *HPS3* (Hermansky-Pudlak Syndrome 3 Protein, biogenesis of lysosomal organelle complex 2 subunit 1). b) Box plots representing expression levels (normalized expression count) of one differentially expressed targeted gene according to interaction term (diagnosis and genetics interaction). Disease and genetic status are labeled on the x-axes. Boxes are colored based on disease and genetic status. Each dot represents a subject. Dots are colored based on *GBA* mutations (as reported in the legend: *GBA* mild mutations (N370, E326K, R496H), *GBA* severe mutations (L444P/A456P/RecNciI, V394L, 84GG, 84GG/T369M, N370S/RecNciI)). p-value of different expression levels is reported on top (statistics: Mann-Whitney U test). Asterisks indicate significant p-value (* = p-value < 0.05, ** = p-value < 0.01, *** = p-value < 0.001).

**Supplementary Fig. 8.**
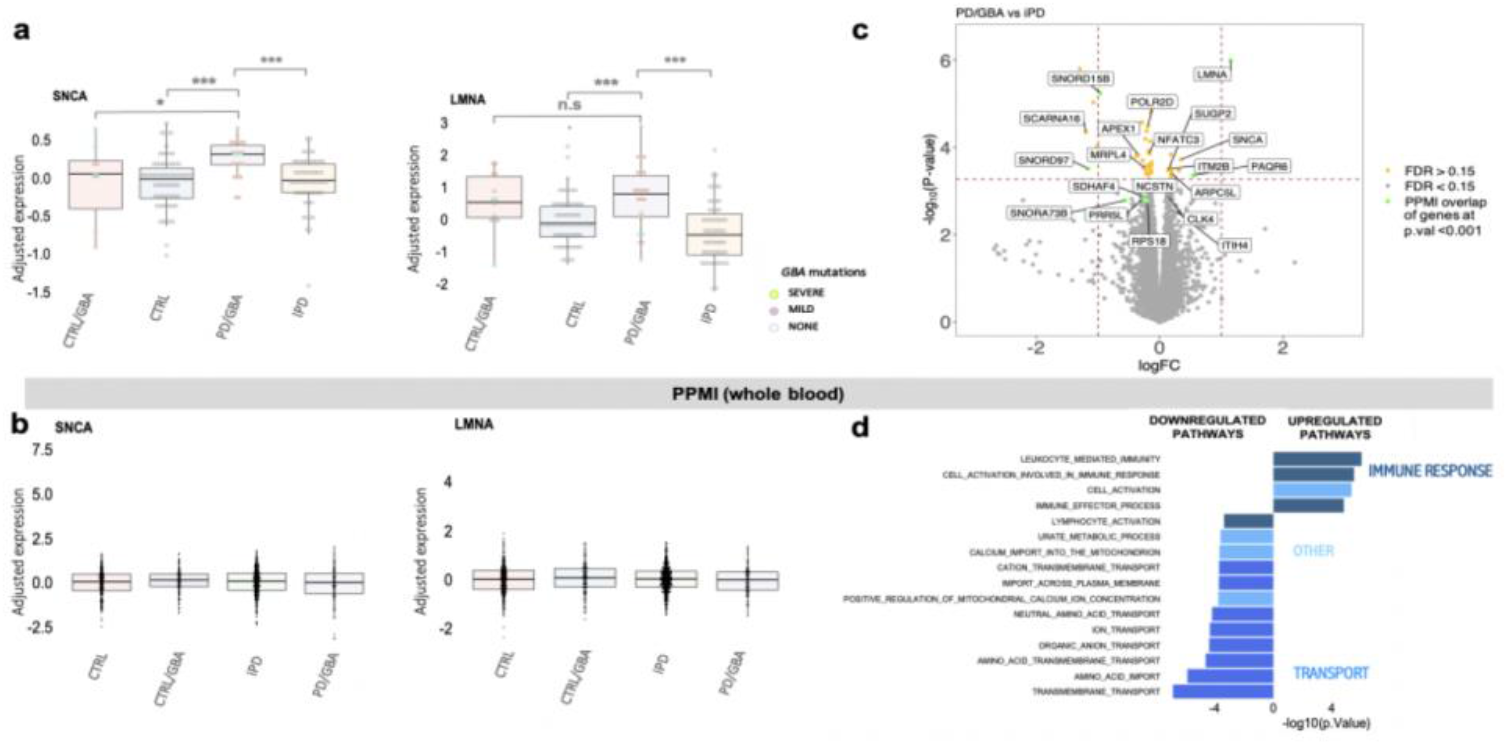
Differential expression analysis in whole blood and monocytes in PD patients with and without *GBA*-variants. a-b) Differential normalized expression count of *SNCA* and *LMNA* between PD/GBA and iPD, compared to CTRL/GBA and CTRL subjects in isolated CD 14+ monocytes (a) and in whole blood (b). Asterisks indicate significant p-value (* = p-value < 0.05, ** = p-value < 0.01, *** = p-value < 0.001). Disease and genetic status are reported on the x-axes. Each dot represents a subject. Dots are colored based on *GBA* mutations (as reported in the legend: *GBA* mild mutations (N370, E326K, R496H), *GBA* severe mutations (L444P/A456P/RecNciI, V394L, 84GG, 84GG/T369M, N370S/RecNciI)). p-value of different expression levels is reported on top (statistics: Mann-Whitney U test). c) Volcano-plot representing logFC (x-axes) and p-value (y-axes, -log_10_ p-value) of differential expressed genes between PD/GBA and iPD as per nested interaction model. Highlighted in yellow are genes with FDR < 0.15 (44 total genes). ID labels of functionally relevant genes and of genes differentially expressed in whole blood (PPMI overlap of genes at nominal p-value <0.001) are reported in the plot. d) Pathway enrichment analysis of differentially expressed genes in whole blood between PD/GBA vs iPD subjects with p-value < 0.01 for GO terms are reported.

**Supplementary Figure 9.**
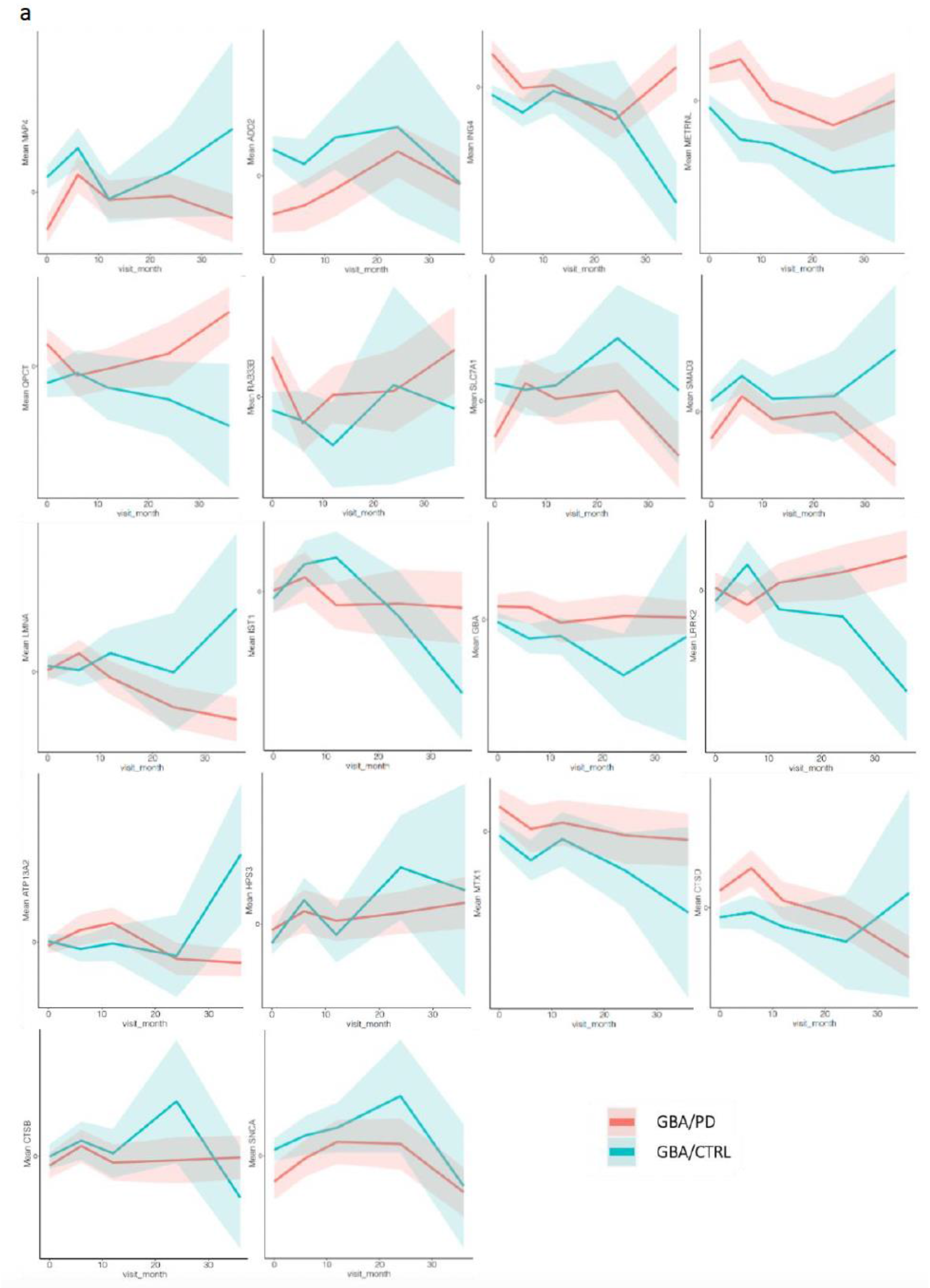
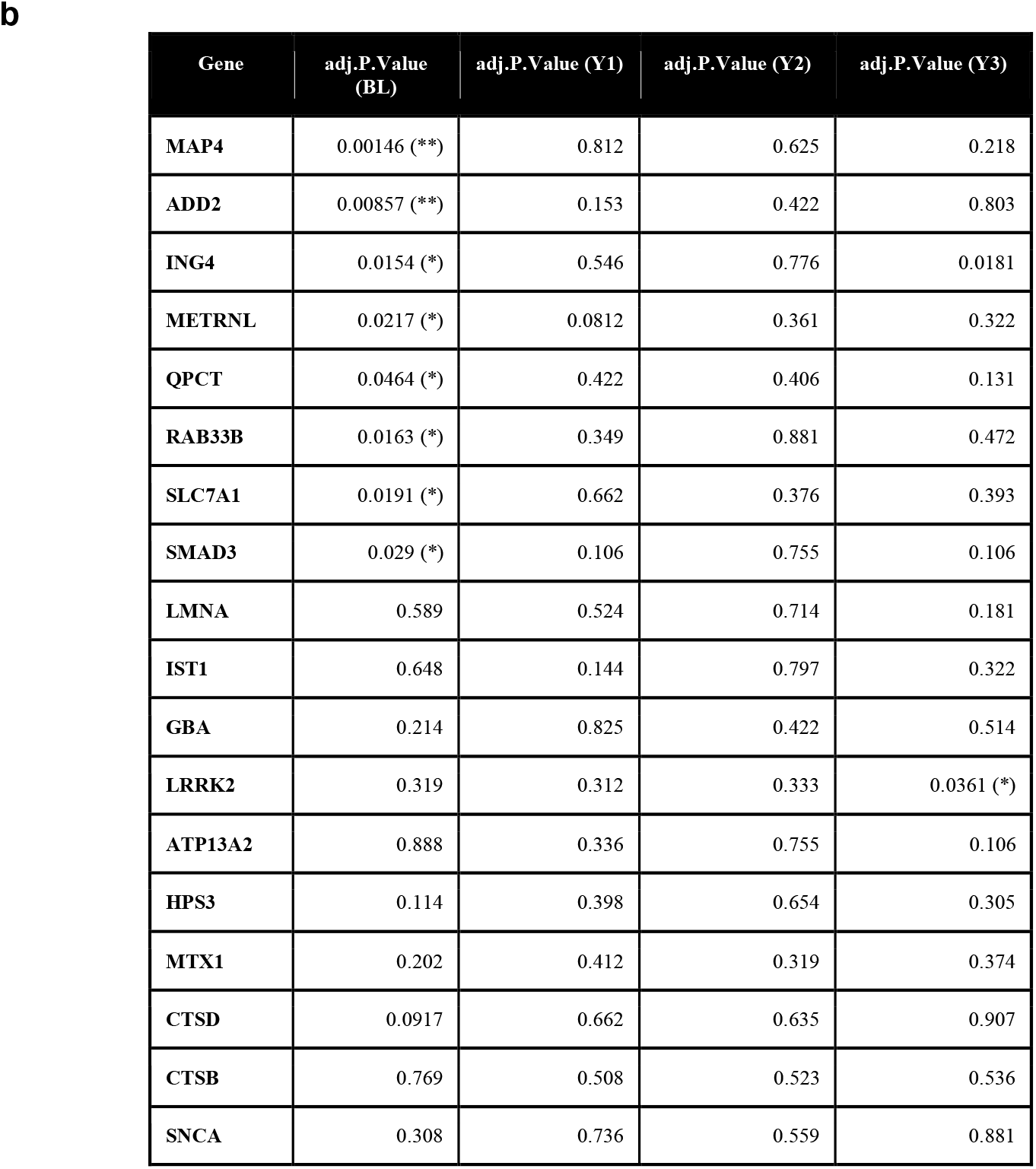
Longitudinal expression profiles in whole blood of targeted genes. a) Data represent normalized expression levels of selected genes across three year follow-up data in manifesting vs non-manifesting carriers (the different cohorts (PD/GBA vs, CTRL/GBA, iPD, CTRL). Data are expressed as mean and confidence interval at 95% (shadow). b) Adjusted p.value was calculated between the two groups at each time point with wilcox_test followed by Bonferroni test and results are reported in the table. Number of subjects in the two groups across visits are as follow: baseline (BL): GBA/PD 122 vs CTRL/GBA 164; Year 1 (Y1): GBA/PD 52 vs CTRL/GBA 37; Year 2 (Y2): GBA/PD 40 vs CTRL/GBA 8; Year 3 (Y3): GBA/PD 38 vs CTRL/GBA 6. Asterisks indicate significant p-value (* = p-value < 0.05, ** = p-value < 0.01, *** = p-value < 0.001).

**Supplementary Fig. 10.**
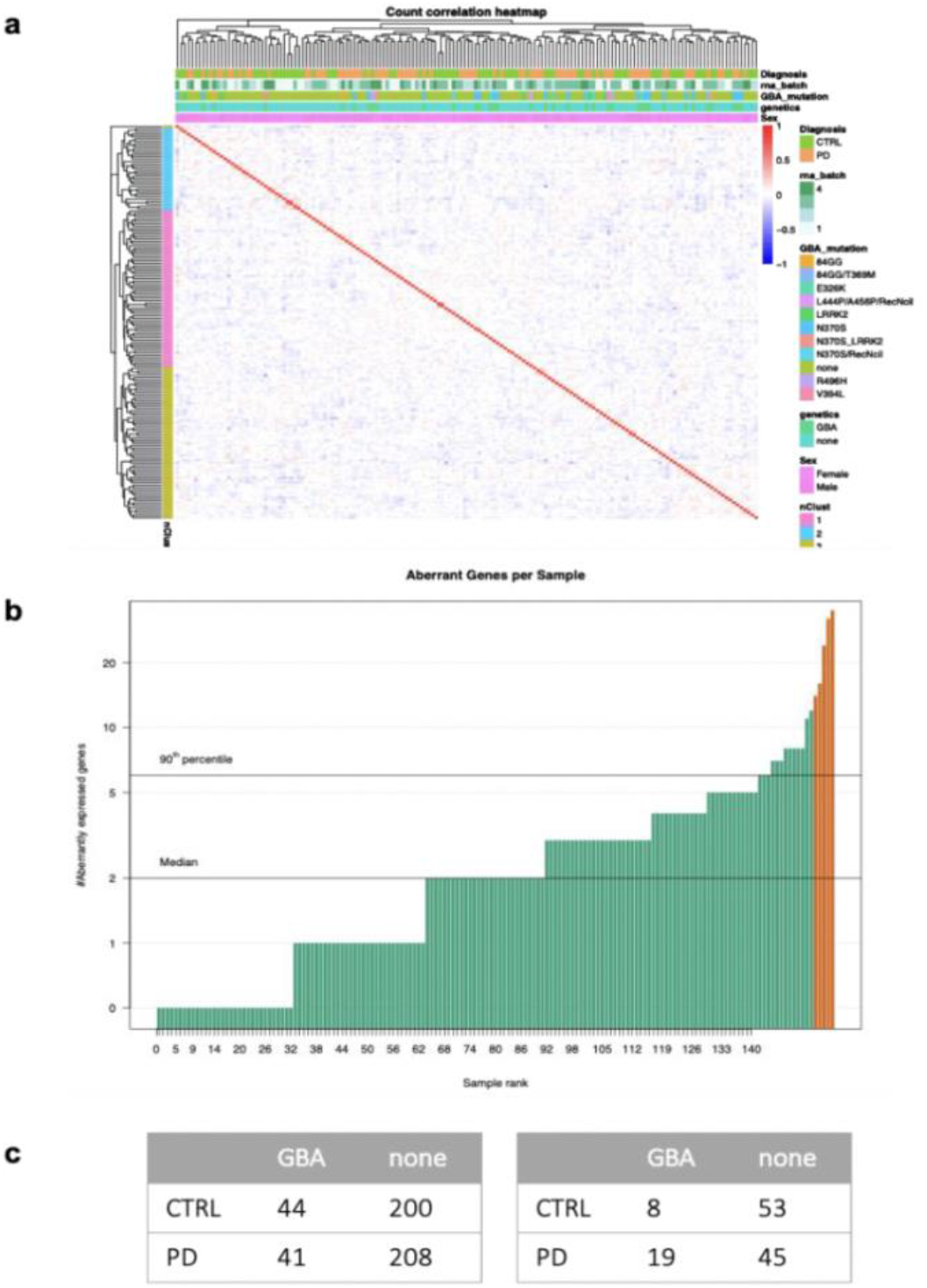
QC for analysis of outlier genes. a) Normalization based on surrogate variables, as provided by the OUTRIDER script, of a total of 13711 genes (considering only genes with > 30% of total expression). Discrete relevant variables (Diagnosis, batches of RNAseq analysis (rna_batch), GBA mutations and GBA-related genetic status (carriers or non-carriers of GBA mutations), gender (Sex: male (M) and female (F)) are labeled at the top of the heatmap per each subject. b) Bar-plot reporting number of outlier genes per each subject (out of 158 subjects). Highlighted in orange samples with outlier gene count above 0.1%. c) Summary tables: on the left: number of outliers genes per cohort (PD/GBA, CTRL/GBA, iPD, CTRL) (493 pairs); on the right: number of subjects per each cohort with at least one outlier gene (125 unique subjects total with at least one outlier gene).

**Supplementary Fig. 11.**
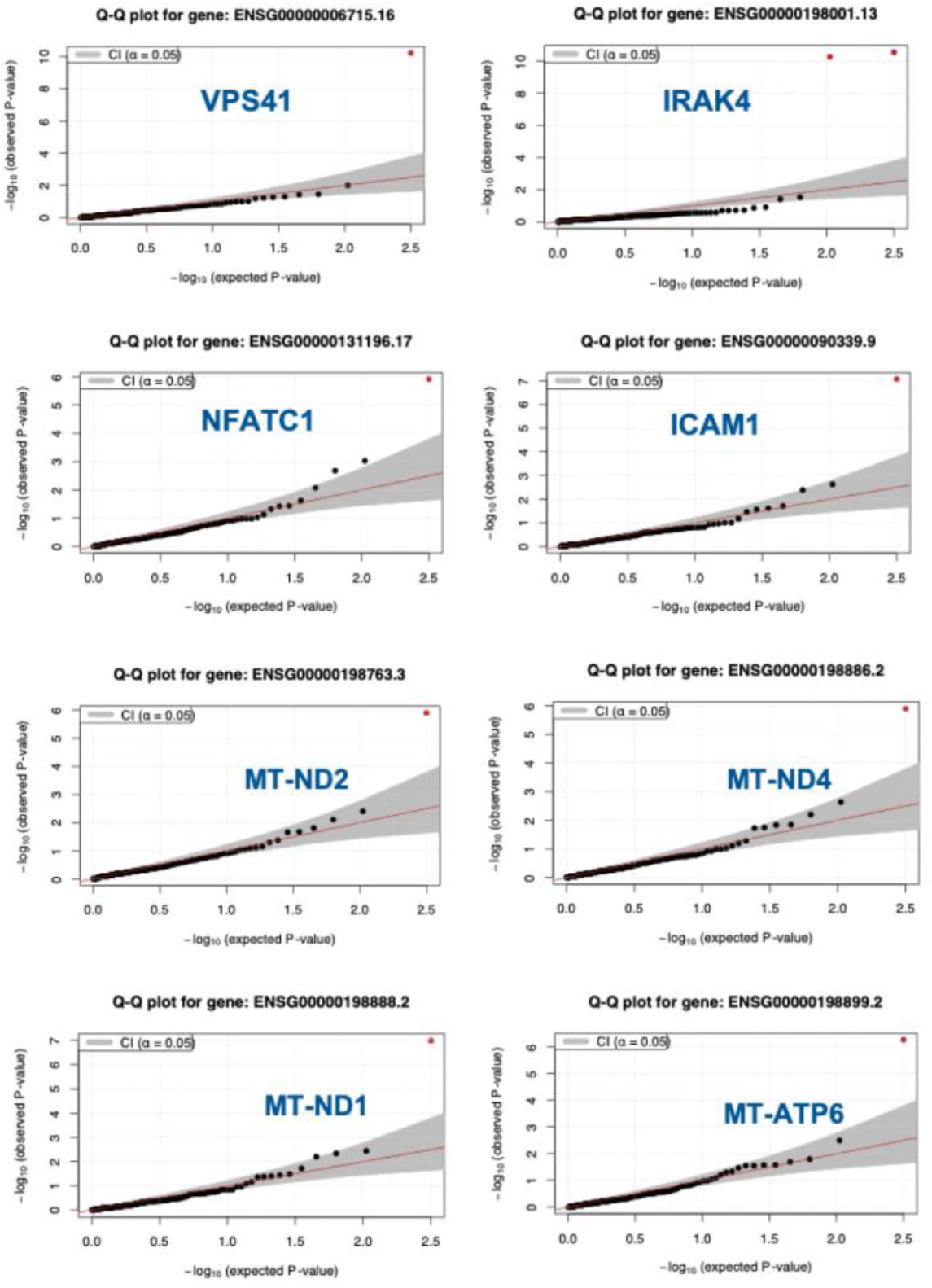
Scatter plot of selected genes identified through the OUTRIDER method. Scatter plot representing -log_10_ (p-value) of outliers genes identified in previous analysis.

**Supplementary Table 1.** Clinical characterization of study cohort. Summary of demographic, clinical and genetic features of the cohort of subjects (PD and CTRL) whose purified CD14+ monocytes were used for integrated genomic analysis.

|  | iPD | PD/GBA | CTRL | CTRL/GBA |
| --- | --- | --- | --- | --- |
| Number of subjects | 56 | 23 | 66 | 13 |
| Gender (% of females) | 30% | 61% | 67% | 54% |
| Race (% of self-reported Caucasians) | 100% | 100% | 100% | 100% |
| Age (mean, min-max) | 68.7 (47-88) | 60.3 (28-77) | 67 (37-86) | 58.3 (38-81) |
| % of N370 GBA-variant carriers | 0 | 74% | 0 | 46% |
| % of G2019S LRRK2-variants carriers | 7% | 4.3% | 1.5% | 0 |

**Supplementary Table 2.**
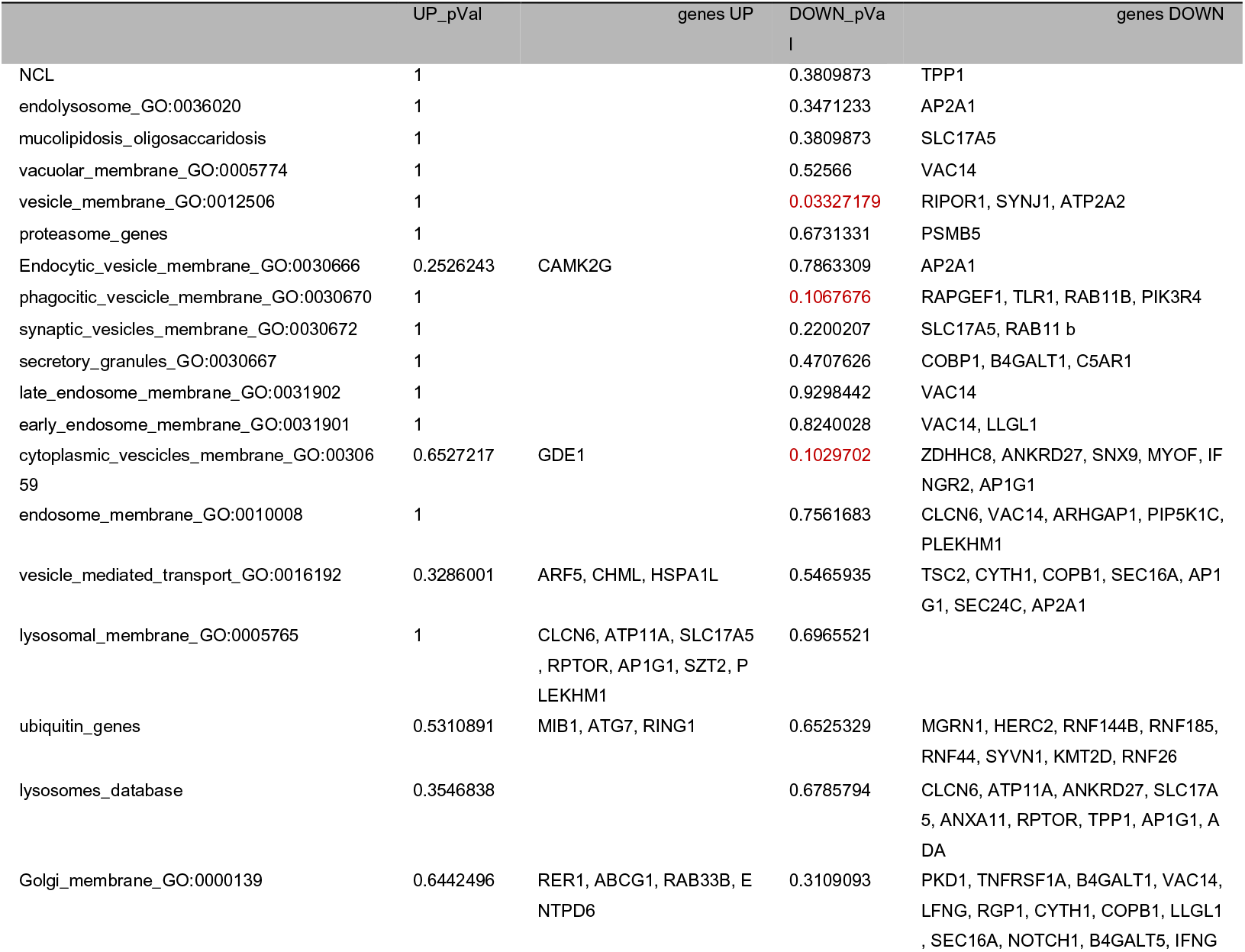
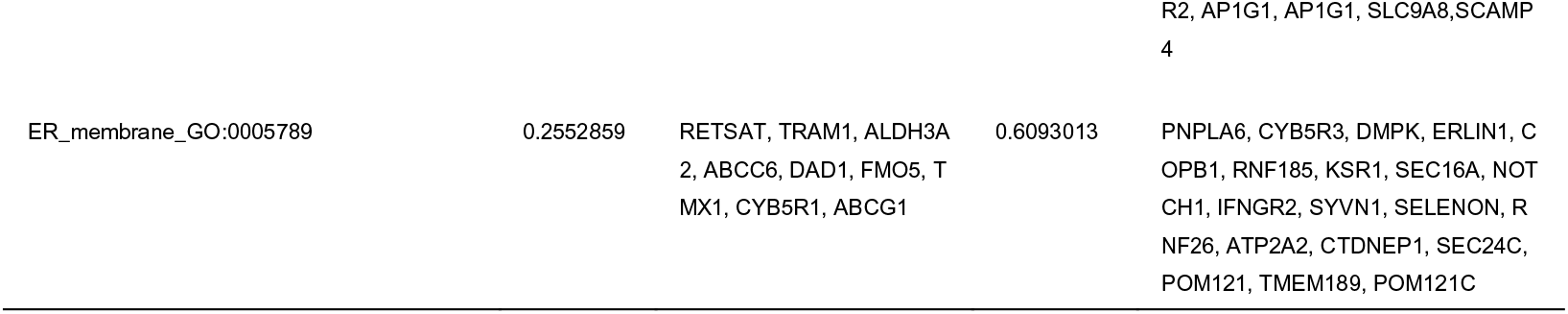
Targeted pathway enrichment in PD/GBA vs CTRL/GBA. List of endolysosomal pathways (from GO terms and curated pathways, i.e. ubiquitin pathway) reported in Figure 6. p-value of enrichment as per Fisher exact test of each pathway within the set of up-regulated (UP) and down-regulated (DOWN) genes. Significant enriched pathways (at p-value < 0.15) are highlighted in red.

**Supplementary Table 3.** GO terms pathways enrichment analysis. The table reports the GO terms and ID for the pathways that were tested for enrichment analysis of differentially expressed genes between the CTRL/GBA and PD/GBA cohorts.

| GO term | GO ID |
| --- | --- |
| vesicle-mediated transport | GO:0016192 |
| endolysosome | GO:0036020 |
| lysosomal membrane | GO:0005765 |
| vacuolar membrane | GO:0005774 |
| mitochondrial membrane | GO:0031966 |
| Golgi membrane | GO:0000139 |
| autophagosome membrane | GO:0000421 |
| vesicle membrane | GO:0012506 |
| endosome membrane | GO:0010008 |
| membrane invagination | GO:0010324 |
| membrane biogenesis | GO:0044091 |
| membrane assembly | GO:0071709 |
| membrane docking | GO:0022406 |
| phagocytic vesicles membrane | GO:0030670 |
| ER membrane | GO:0005789 |
| peroxisomal membrane transport | GO:0015919 |
| mitochondrial membrane fusion | GO:1990613 |
| synaptic vesicles membrane | GO:0030672 |
| secretory granule membrane | GO:0030667 |
| mitochondria outer membrane | GO:0005741 |
| mitochondrial inner membrane | GO:0005743 |
| coated vesicles membrane | GO:0030662 |
| endocytic vesicles membrane | GO:0030666 |
| cytoplasmic vesicles membrane | GO:0030659 |
| transport vesicles membrane | GO:0030658 |
| autophagosome membrane docking | GO:0016240 |
| peroxisome membrane biogenesis | GO:0016557 |
| exocytic vesicles membrane | GO:009950 |
| mitochondrial membrane fission | GO:0090149 |
| early endosome membrane | GO:003190 |
| late endosome membrane | GO:0031902 |
| early phagosome membrane | GO:0036186 |
| complement activation | GO:0006956 |
| extracellular exosomes: | GO:0070062 |
| exosomes (RNA complex) | GO:0000178 |
| extracellular exosomes complex | GO:1990563 |
| extracellular exosomes biogenesis | GO:0097734 |
| extracellular exosomes micropinocytosis | GO:0061707 |
| extracellular exosomes assembly | GO:0071971 |
| cytoplasmic exosomes | GO:0000177 |
| trans-synaptic signaling via exosomes | GO:0099157 |
| regulation of extracellular exosomes assembly | GO:1903551 |
| clathrin dependent exosomes assembly | GO:1990771 |
| positive regulator of exosome assembly | GO:1903553 |
| negative regulator of exosome assembly | GO:1903552 |
| RNA polymerase exosomes dependent | GO:0030847 |
| exosomal secretion | GO:1990182 |
| regulation of exosomal secretion | GO:1903541 |
| negative regulation exosomal secretion | GO:1903542 |
| positive regulation exosomal secretion | GO:1903543 |

**Supplementary Table 4.** Overlapping genes between monocytes and PPMI cohort (whole blood) in manifesting vs non-manifesting GBA-carriers. List of overlapping genes differentially expressed in monocytes and whole blood in manifesting vs non-manifesting carriers. Gene name, description of gene function and logFC and FDR in monocytes and whole blood are reported. In “grey” genes that represent PD-GWAS hits are highlighted.

| Gene | Description | logFC<br>(monocytes) | FDR<br>(monocytes) | logFC<br>(whole blood) | FDR<br>(whole blood) |
| --- | --- | --- | --- | --- | --- |
| **ABCG1** | Phospholipids and cholesterol cellular transport | 0.961465911314694 | 0.0459624608087888 | 0.23968699380249 | 0.0401794415274729 |
| **AC007342.3** |  | 0.57210658223937 | 0.0116606709179357 | 0.283178419304563 | 0.00823687604203322 |
| **ADD2** | β adducin gene / assembly of the spectrin-actin network | -0.912023451806256 | 0.0483997995924918 | -0.141246049132182 | 0.0272036288413351 |
| **ARHGAP9** | Rho GTPase cycle, Neutrophil degranulation, hematopoietic cells binding to extracellular matrix | 0.268124153126283 | 0.0101875885983114 | 0.235030480304674 | 0.0182020701710991 |
| **GPI** | Glucose-6-Phosphate Isomerase / lipid anchor for cell-surface proteins | -0.186214271347422 | 0.0400440200555539 | -0.143507177670195 | 0.0244152975439078 |
| **ING4** | Inhibitor Of Growth Family Member 4 / tumor suppressor / apoptosis | 0.198598151130411 | 0.0464557273237816 | 0.0916280556322153 | 0.0191529717760595 |
| **MAP4** | Microtubule-associated protein 4 /microtubule associated protein related to TAU and STMN1 (GWAS hit) | -0.322611615992687 | 0.0170722305590492 | -0.149920509213461 | 0.0113155729337565 |
| **METRNL** | Meteorin-like/ expressed in activated macrophages | -0.376611923492884 | 0.0348741627170725 | 0.158803045181993 | 0.0354498086303043 |
| **MTHFD1** | methylenetetrahydrofolate dehydrogenase 1 | -0.293226296096193 | 0.0459624608087888 | -0.137985273190468 | 0.0464314181999015 |
| **QPCT** | Glutaminy-peptide cyclotransferase / metabolism APP amyloid-beta peptides | 0.294663529079197 | 0.035839785633954 | -0.0017683378784259 | 0.98649984057878 |
| **RAB33B** | Ras-related protein Rab-33B / autophagosome formation / Golgi homeostasis and trafficking | 0.272912365448382 | 0.0236446424550435 | 0.141779591452484 | 0.0273850778831599 |
| **RGL2** | Ral guanine nucleotide dissociation stimulator-like 2 (GWAS hit) | 0.177237657505668 | 0.0348741627170725 | 0.177270448436024 | 0.0335312353454597 |
| **RSRP1** | Arginine And Serine Rich Protein 1 | 0.294899104257617 | 0.016281087770457 | 0.109410987627835 | 0.0267370496570794 |
| **SLC7A1** | High affinity cationic amino acid transporter 1 | -0.395917432544339 | 0.0363324920815811 | -0.255699529915818 | 0.0370438163669751 |
| **SMAD3** | TGF-beta signaling pathway | -0.456474255965785 | 0.0156414130224551 | -0.168543588997348 | 0.0169091684342028 |
| **SUPV3L1** | ATP-dependent RNA helicase SUPV3L1, mitochondrial/ maintenance of mtDNA and chromatin | -0.332545474992509 | 0.0101676754748312 | -0.0859904726039504 | 0.0172965956631605 |

